# Fine-mapping SLE-MHC associations revealed independent contributions of HLA missense variants and *C4* copy number variations

**DOI:** 10.1101/2024.11.21.24317596

**Authors:** Chae-Yeon Yu, Dong Mun Shin, Sung Min Kim, Yui Taek Lee, Sungwon Jeon, Sehwan Chun, So-Young Bang, Hye-Soon Lee, Xianyong Yin, Yong Cui, Xuejun Zhang, Jong Bhak, Soon Ji Yoo, Young Jin Kim, Bong-Jo Kim, Sang-Cheol Bae, Kwangwoo Kim

## Abstract

Systemic lupus erythematosus (SLE) is a complex autoimmune disease with strong genetic associations within the major histocompatibility complex (MHC) region. Despite significant advances, precisely pinpointing the genetic variants that contribute to SLE risk within the MHC remains challenging. This study aimed to comprehensively profile SLE-driving variants using a newly developed East Asian MHC imputation reference panel, capable of simultaneously imputing diverse MHC variants, including multi-level variants of HLA genes and copy number variations (CNVs) of *C4* elements, with high imputation accuracy. Applying this panel to two SLE genome-wide association study datasets, we uncovered the independent contributions from six amino acid positions altering the epitope-binding surfaces of HLA-DRB1 and HLA-C. Additionally, reduced *C4A* copy numbers and increased HERV copy numbers, collectively lowering C4 protein levels, were associated with increased SLE risk, independent of HLA variants. Our refined MHC-SLE association model provided superior explanations for SLE risk over previous association models. In summary, this study enhanced the understanding of HLA and *C4* in SLE pathogenesis and holds promise for advancing MHC association studies for immune-mediated inflammatory disorders in East Asians using our MHC panel (https://coda.nih.go.kr/usab/kis/intro.do).

## Introduction

Systemic lupus erythematosus (SLE) is a common polygenic autoimmune disorder characterized by complex dysregulations in both adaptive and innate immune systems, resulting in the breakdown of self-tolerance and impaired clearance of nuclear antigens^1^. As of 2024, genome-wide association studies (GWASs) have identified over 200 loci in the human genome associated with susceptibility to SLE^2,3^. Most of these associations are driven by single genetic signals, with several notable exceptions such as the major histocompatibility complex (MHC) region, which exerts particularly strong effects on SLE risk^2^.

The MHC region on human chromosome 6 harbors a diverse array of immune-related genes, featuring human leukocyte antigens (HLAs), and complements, and cytokines. This region has been well documented for its extensive linkage disequilibrium (LD), exceptionally dense distribution of genetic variants, and the presence of copy number variations (CNVs). These factors contribute to the formation of various long-range haplotypes across the MHC region, including HLA classical alleles and other MHC genes. The resulting complex array of haplotypic combinations influences the functional diversity of immune system and susceptibility to various diseases. Additionally, this intricate genomic architecture has made the MHC region one of the most challenging loci to characterize in terms of individuals’ genetic variants and to dissect genetic association signals^4^.

Our previous association fine-mapping studies of the MHC region have demonstrated the crucial role of structural changes in the epitope-binding groove of HLA-DR molecules in multiple Asian populations^5–7^. These changes are determined by HLA-DRB1 missense variants encoding amino acid positions 11, 13, 26, and 37. The haplotypes of residues at these positions accounted for the largest proportion of the MHC association effects on SLE risk, explaining both the previously reported associations of *HLA-DRB1* classical alleles and the association heterogeneity between East Asians and Europeans, such as the association of *HLA-DRB1*15:01* in Asians and *HLA-DRB1*03:01* in Europeans^5^.

In parallel, some studies have reported genetic associations of the complement component gene *C4*, located near *HLA-DRB1*, with SLE. *C4* is crucial in the human immune system, facilitating the clearance of dead or damaged cell debris. The genetic variants of *C4* are highly complex, involving various combinations of coding variants, endogenous retroviral sequence insertions, and CNVs. *C4* encodes two functionally distinct isoforms, *C4A* and *C4B*, which differ by four amino acid residues within positions 1101 to 1106 and exhibit differential affinities for their molecular targets associated with different biological pathways^8,9^. Additionally, the insertion of human endogenous retrovirus (HERV) segments in intron 9 of *C4A* or *C4B* is common in the general population. The HERV-inserted form, often referred to as the long form, contrasts with the short form and is associated with decreased *C4* expression in blood^10^. Most importantly, the total copy number of *C4* varies widely among individuals, mostly ranging from two to eight copies, consisting of various copy numbers of both *C4A* and *C4B*, with and without HERV segments. Therefore, the variance in expression levels of the functionally distinct C4A and C4B isoforms in a general population is attributed mainly to the CNV of *C4*, with additional modulation by the HERV insertion^9^. The complex genomic architecture of *C4* requires relatively low-throughput, expensive technologies to characterize individuals’ genetic variants. As a result, most *C4*-SLE association studies have been conducted with relatively small sample sizes^11–17^. Furthermore, due to the lack of an imputation reference panel that simultaneously infer both HLA and *C4* variants along with other MHC variants, previous *C4* genetic association studies have focused solely on the effects of the *C4* CNV on SLE, without considering the compounding effects of HLA variants^11–17^.

A recent study by Kamitaki *et al.,* reported a primary MHC-SLE association signal at *C4*, rather than *HLA-DRB1*03:01* using data from large European and African ancestral populations^18^. Although the primary genetic effect could not be distinctly attributed to either *C4* CNVs or *HLA-DRB1*03:01* in European ancestry due to relatively high LD, a joint logistic regression analysis for *C4* CNVs and *HLA-DRB1*03:01* in African Americans revealed a sole association of *C4* CNVs with SLE, while *HLA- DRB1*03:01* showed no significant association. This finding challenges the long-held belief regarding *HLA-DRB1* associations with SLE, suggesting that the SLE-MHC association, traditionally attributed to *HLA-DRB1*, may instead be driven by CNVs of *C4* alleles.

However, we urge cautious interpretation of these results for several reasons. First, the study reported very poor imputation accuracy in *C4* copy number calls among African populations, potentially impacting the reliability of the findings. Second, the most significant variant was not mapped to *C4* in either ancestry. In an unconditional analysis, the study observed a stronger significant association of *HLA-DRB1*03:01* compared to the *C4* CNV in Europeans. Similarly, another recent European study identified *HLA-DQB*02:01* as the most significant variant for SLE rather than *C4* CNVs^19^. In Africans, *HLA-DRB1*15:03*, known as the most significant classical allele in this population, also showed a stronger association with SLE compared to the *C4* CNVs, in their study and others^20,21^. Third, adjusting solely for *HLA-DRB1*03:01* may not fully capture the multi-allelic nature of *HLA-DRB1* associations. Residual effects after conditioning on *HLA-DRB1*03:01* are expected to remain substantial around and within *HLA-DRB1*. Furthermore, in African ancestry, *HLA- DRB*03:01* contributes much less to the risk of SLE.

In our present study, to better understand the MHC associations in SLE through a sophisticated, comprehensive genetic analysis using large-scale case-control GWAS data, we have developed an innovative MHC imputation reference panel from whole-genome sequencing (WGS) data by analyzing various MHC variants, including *C4* and HLA genes, and phasing them into haplotypes. Unlike existing MHC reference panels, our panel allows for the simultaneous imputation of copy numbers for *C4A*, *C4B*, and HERV, as well as amino acid residues and classical alleles of eight major HLA genes, along with neighboring SNPs and indels across the MHC region. The imputation performance of our reference panel has been assessed through various approaches, achieving a high level of accuracy. Applying our imputation reference panel to large SLE GWAS datasets, we uncovered multiple independent genetic contributions from *HLA-DRB1*, *C4*, and *HLA-C*. This led us to propose a novel SLE-MHC association model that, for the first time, incorporates HLA missense variants at *HLA-DRB1* and *HLA-C*, and the copy numbers of *C4A* and HERV. The analysis schematic of this study is illustrated in **Fig. 1**.

**Fig. 1.**
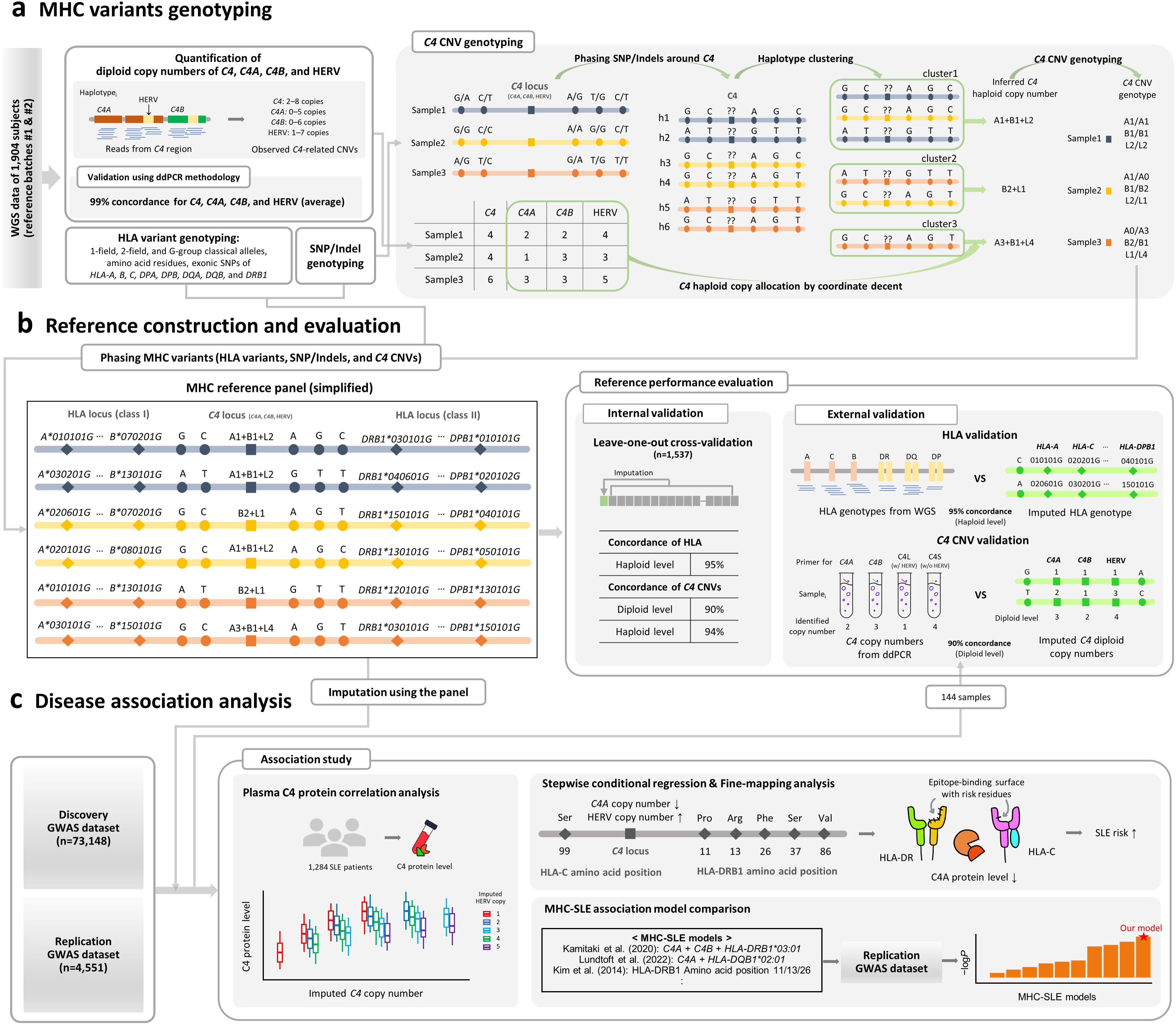
Overview of the study design. The schematic diagram illustrates the key analysis steps of the study. **a.** To construct an MHC imputation reference panel, WGS data from 1,904 samples in two independent batches were used to genotype classical alleles and amino acid residues in eight major HLA genes, CNVs of *C4*-related elements, and other MHC SNPs/indels. For *C4*, the WGS-based quantification of copy numbers was performed at the diploid level, and the reliability of this quantification method was further validated using ddPCR. The diploid copy numbers of *C4A*, *C4B*, and HERV were then segregated into haploid copy numbers through unsupervised haplotype clustering and by determining the most likely haploid copy number of each *C4*-related element using a cyclic coordinate descent method, based on the WGS-based diploid copy numbers across the reference individuals. For instance, the example cluster “cluster1” comprises three haplotypes that are likely to possess a single copy of *C4A*, a single copy of *C4B*, and two copies of HERV (designated as A1+B1+L2). **b.** The MHC imputation reference panel was constructed by phasing all binary-coded MHC variants into haplotypes. Its imputation performance was evaluated based on the concordance rates between actual and imputed data from both reference and external samples. **c.** The newly developed imputation reference panel was applied to two independent SLE GWAS datasets: discovery and replication datasets. Plasma C4 protein levels measured in 1,284 SLE patients were analyzed for correlations with imputed *C4*-related copy numbers, revealing a positive correlation with *C4* copy number and a negative correlation with HERV copy number. Finally, through stepwise conditional and fine-mapping analyses, three independent signals were identified at *HLA-DRB1*, *C4*, and *HLA-C*, which included key variants contributing to the risk of SLE: amino acid positions 11, 13, 26, 37, and 86 in the epitope-binding pocket of HLA-DRB1, position 99 in the epitope-binding pocket of HLA-C, and the copy numbers of *C4A* and HERV in *C4*. Our refined MHC association model was compared with previous models using the replication dataset, evaluating model significance to confirm that the variants identified through our fine-mapping analysis provided the most robust explanation for SLE risk.

## Results

### Development of the MHC reference panel for HLA and *C4* imputation

The MHC reference panel for imputing diverse MHC variants, including HLA and *C4* variants, was constructed to include >3,000 haplotypes of the general Korean population, based on WGS data from two independent experimental batches. To achieve this, we genotyped classical alleles of eight major HLA genes (*HLA-A*, *-B*, *-C*, *-DPA1*, *-DPB1*, *-DQA1*, *-DQB1*, and *-DRB1*) at the G-group resolution using a highly accurate graph-based genotyping method, HLA-LA^22^ (**Supplementary Fig. 1**). We further defined these alleles at one-field and two-field resolutions and retrieved their amino acid residue information. In our reference samples, we observed 102 classical alleles at one-field resolution, 267 at two-field resolution, and 289 at G-group resolution, along with 1,440 polymorphic amino acid positions.

The diploid copy numbers of *C4*, *C4A*, *C4B*, and HERV were estimated using Genome STRiP^23^, based on read depth and the allele-specific read ratio between *C4A* and *C4B*, as detailed in **Methods** (**Supplementary Fig. 2**). We observed a wide range of diploid gene copy numbers of *C4*, from 2 to 8 in the reference samples. Similarly, the diploid copy number of the allele-specific isoforms of *C4* ranged from 0 to 5 for *C4A*, and from 0 to 6 for *C4B*. The HERV segment within *C4* was detected with diploid copy numbers ranging from 1 to 7 copies (**Supplementary Fig. 3**). The diverse combinations among CNVs, alleles, and HERV inserts greatly expanded the genetic variability of *C4* (**Fig. 2a**). For instance, the most common combination was to carry four copies of *C4*, which included two *C4A* copies, two *C4B* copies, and at least two copies of HERV.

**Fig. 2.**
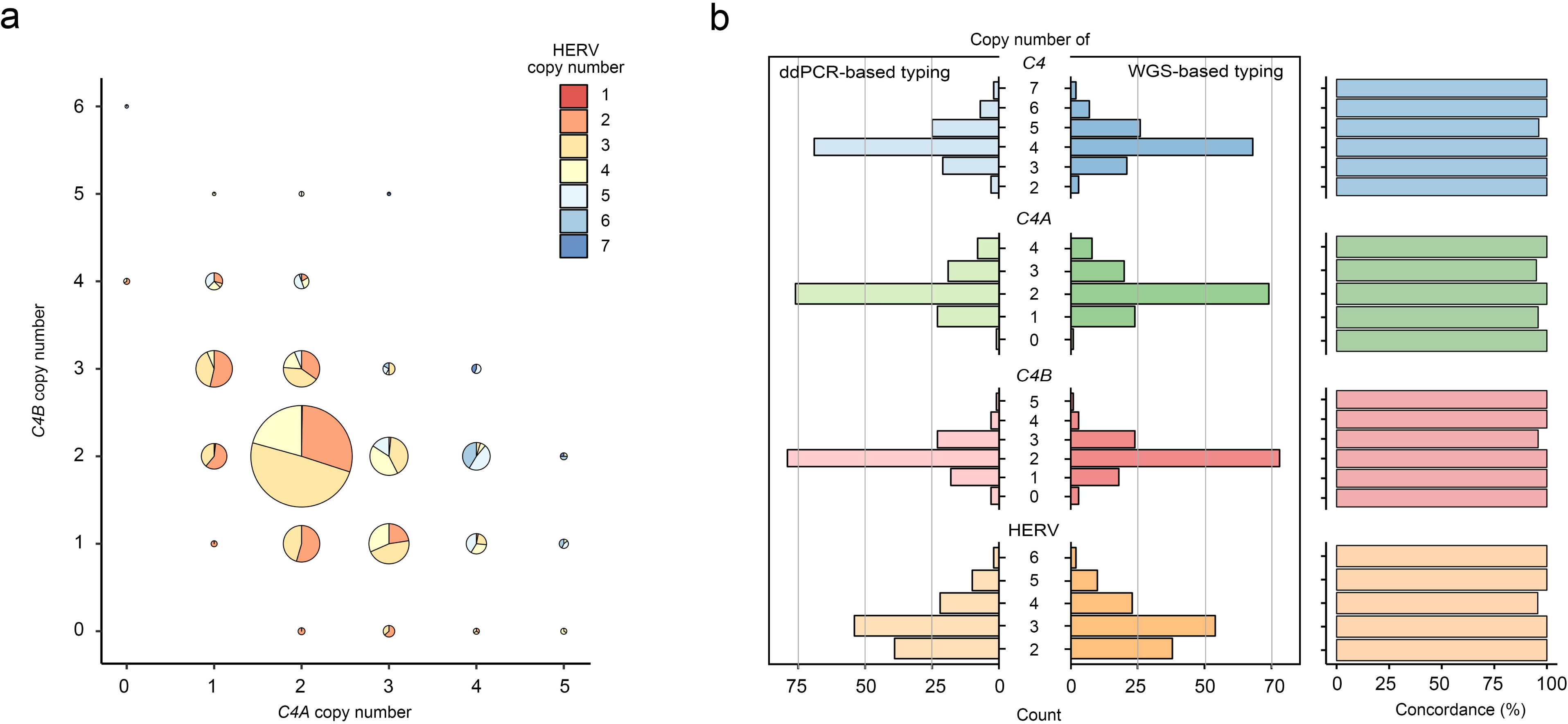
Distribution of diploid copy numbers of *C4*. **a.** The joint distribution of diploid copy numbers of *C4A*, *C4B*, and HERV was examined using WGS data for reference samples (n=1,537). The size of each data point is proportional to the frequency of samples with the corresponding combination of *C4A* and *C4B* copy numbers. Each data point is a pie chart illustrating the distribution of HERV copy numbers for the given combination of *C4A* and *C4B* copy numbers. **b.** The reliability of the *C4* copy number quantification method using WGS data was evaluated by comparing WGS-based results with ddPCR-based results from 127 non-reference samples. The bi-directional histogram shows the distribution of diploid copy numbers of *C4*, *C4A*, *C4B*, and HERV, as determined by ddPCR (left) and the WGS-based approach (right). Concordance rates between the two methods are displayed in the bar plot on the right end, according to each copy number of *C4* genes and related elements (≥ 98.4%).

The accuracy of diploid copy number calls for *C4* from WGS data was validated by comparing with those derived from digital droplet polymerase chain reaction (ddPCR) technology^24,25^ across 127 additional Korean samples (**Supplementary Fig. 4** and **5**). We found highly reliable concordance rates between the WGS-based and ddPCR-based diploid copy numbers (99.2% for *C4*, 98.4% for *C4A*, 99.2% for *C4B*, and 99.2% for HERV) (**Fig. 2b**). Additionally, the distribution of diploid copy numbers observed in this study aligns with previously reported distributions in other populations^16,17,19^.

To segregate each diploid copy number of *C4A*, *C4B*, and HERV into haploid copy numbers, we first defined haplotype clusters based on neighboring SNPs and short indels using unsupervised machine learning, employing various Gaussian mixture variational autoencoder (GMVAE) models^26^ (**Fig. 3a-b**). We then conducted the cyclic coordinate descent method to determine the most likely haploid copy numbers in the linkage with each haplotype cluster, aiming to best explain the observed diploid copy numbers across the entire reference sample set (**Supplementary Table 1** and **Supplementary Fig. 6**). The genotypic combination of possible haploid copy numbers in each sample was determined based on the actual diploid copy number and the likelihood of having each haplotype cluster for each homologous chromosome pair in the sample (**Fig. 3a**). The frequency distributions of determined haploid copy numbers for *C4A*, *C4B*, and HERV showed single-modal patterns, each centered around one copy, and this pattern was consistently reflected in the copy-number genotype distributions (**Fig. 3c, d**). Stochastically inferred diploid copy numbers from these haplotype frequencies matched the observed diploid copy numbers, indicating that *C4* CNVs followed Hardy-Weinberg equilibrium (HWE) (**Supplementary Table 2**).

**Fig. 3.**
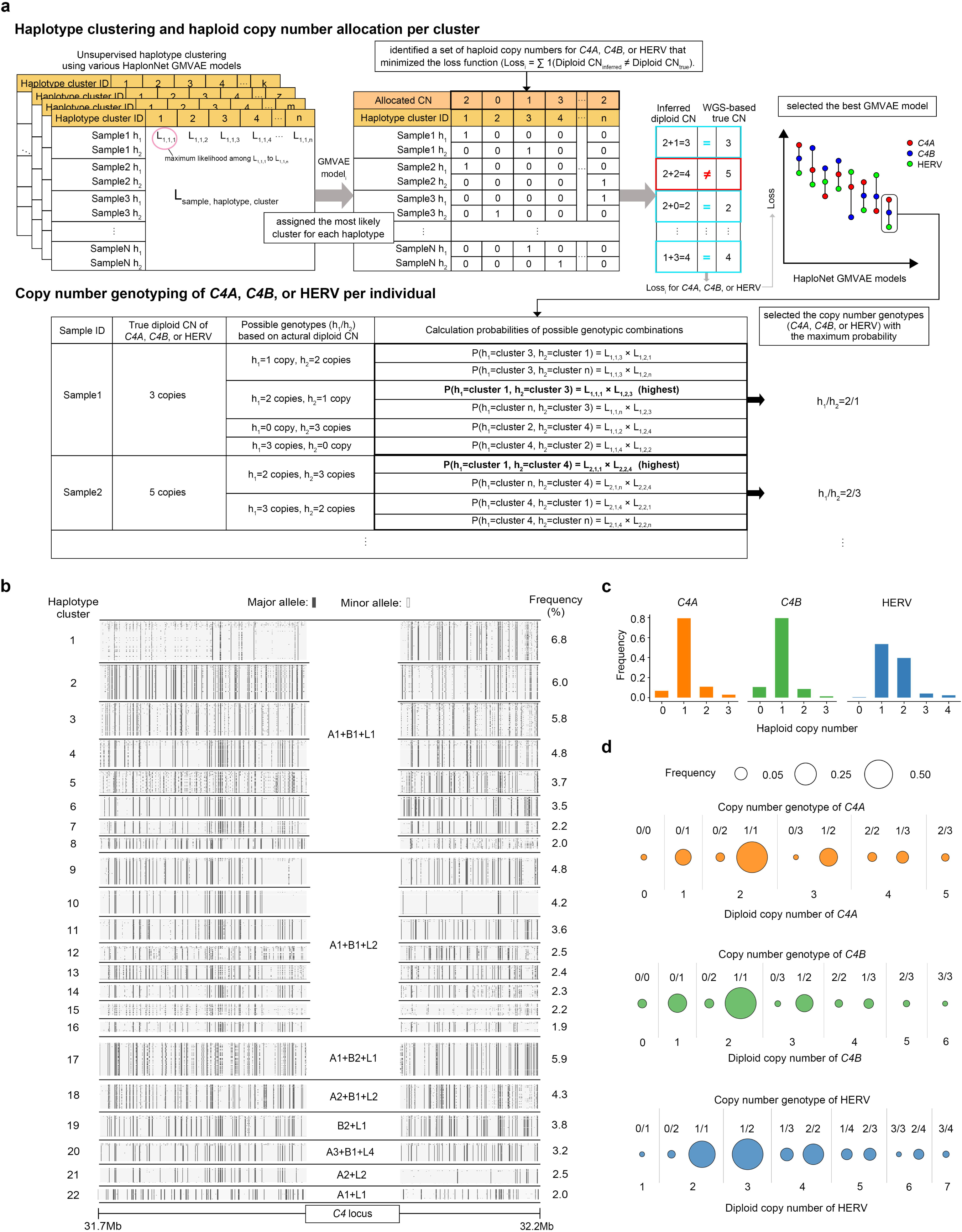
Segregating the diploid copy numbers of *C4A*, *C4B*, and HERV into haploid copy numbers. **a.** Unsupervised haplotype clustering was performed using various GMVAE models based on phased SNPs and indels near *C4* in reference samples, estimating the likelihood of each haplotype belonging to each cluster. For each GMVAE model, each haplotype was assigned to the cluster with the highest likelihood. The most likely haploid copy number for each cluster was then determined using a cyclic coordinate descent method, which iteratively minimized the loss function about the difference between inferred and true diploid copy numbers across reference samples. The GMVAE model with the lowest average loss for *C4A*, *C4B*, and HERV was selected for subsequent procedures. Individuals’ CNV genotypes, consisting of two haploid copy numbers for *C4A*, *C4B*, and HERV, were determined as the most likely haploid copy number combinations, evaluated by multiplying the normalized likelihoods for the two haplotypes belonging to specific clusters, and aligned with the actual diploid copy numbers. **b.** Haplotype clustering analysis near the *C4* region (31.7-32.2Mb on chromosome 6, hg38) identified 47 haplotype clusters. The most common clusters representing 80% of all haplotypes are visualized, along with allocated copy number compositions and frequencies. Each row represents a haplotype, where black and white dots indicate major and minor alleles of bi-allelic variants upstream and downstream of *C4*, respectively. The *C4* composition is labeled using the initials and copy numbers of *C4A* (A), *C4B* (B), and HERV (L; long form). For instance, “A3+B1+L4” denotes the combination of three haploid copies of *C4A*, a single copy of *C4B*, and four copies of HERV (meaning all *C4* copies are in long form. **c.** Distribution of haploid copy numbers for *C4A*, *C4B*, and HERV in reference samples are displayed on the bar plots. **d.** The frequencies of CNV genotypes are plotted according to diploid copy numbers of each *C4*-related elements.

Finally, we assembled the MHC imputation reference panel by incorporating a comprehensive set of 55,632 MHC variants, including HLA variants, *C4*-related CNVs, and other SNPs and indels in 1,537 quality control (QC)-passed samples. These variants were phased into haplotypes, with each allele of multi-allelic variants (e.g., HLA classical alleles, HLA amino acid residues) encoded in a binary format to indicate its absence or presence. Similarly, each haploid copy number of *C4*-related elements (*C4A*, *C4B*, and HERV) was encoded in the same binary format to denote the presence or absence of each possible copy number.

### Performance of the new MHC reference panel in imputing HLA and *C4* variants

We conducted leave-one-out cross-validation, where each reference sample was sequentially excluded, its *C4* and HLA variants were masked, and those variants were then imputed using the MHC reference panel with the remaining reference samples. For HLA genes, our panel demonstrated generally excellent concordance rates across all eight classical HLA genes, including *HLA-DRB1* and *HLA-B*, which have previously shown relatively lower imputation accuracy compared to other HLA genes^27^. The average concordance rates were 99.2% at one-field resolution, 97.6% at two-field resolution, and 97.3% at G-group resolution (**Table 1**).

**Table 1.**
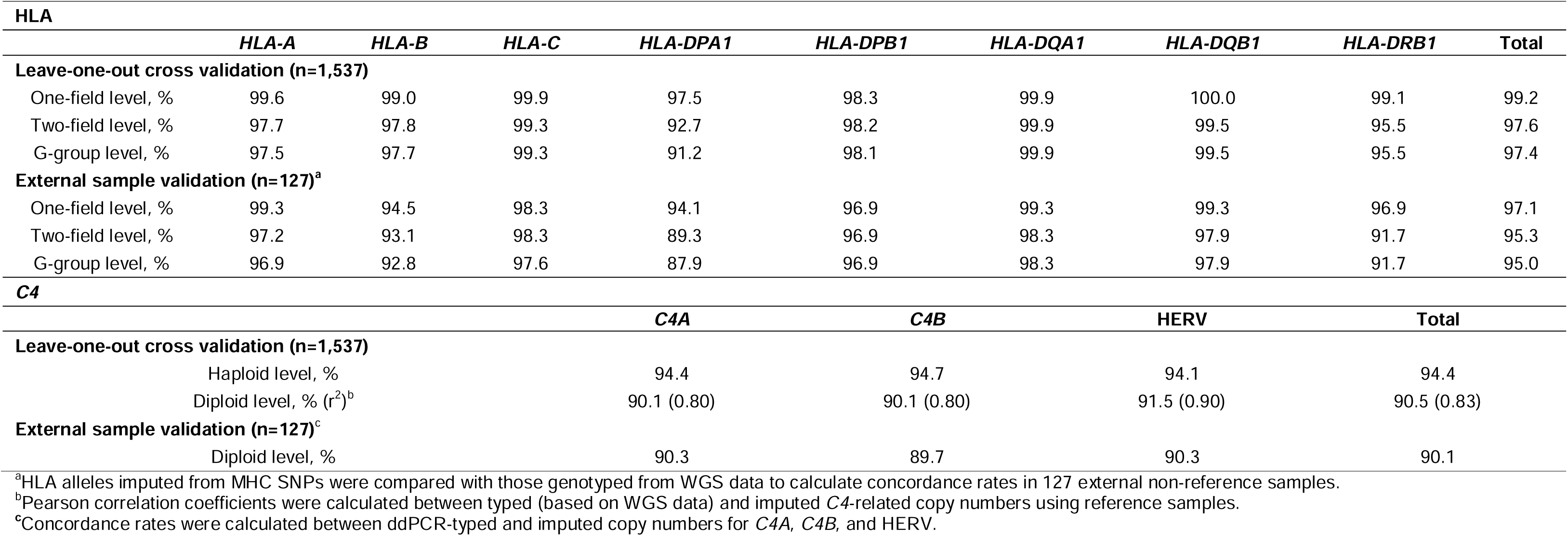
Concordance rates between imputed and actual data for HLA classical alleles and *C4* CNVs from leave-one-out cross validation and external sample validation.

| HLA |  |  |  |  |  |  |  |  |  |
| --- | --- | --- | --- | --- | --- | --- | --- | --- | --- |
|  | HLA-A | HLA-B | HLA-C | HLA-DPA1 | HLA-DPB1 | HLA-DQA1 | HLA-DQB1 | HLA-DRB1 | Total |
| Leave-one-out cross validation (n=1,537) |  |  |  |  |  |  |  |  |  |
| One-field level, % | 99.6 | 99.0 | 99.9 | 97.5 | 98.3 | 99.9 | 100.0 | 99.1 | 99.2 |
| Two-field level, % | 97.7 | 97.8 | 99.3 | 92.7 | 98.2 | 99.9 | 99.5 | 95.5 | 97.6 |
| G-group level, % | 97.5 | 97.7 | 99.3 | 91.2 | 98.1 | 99.9 | 99.5 | 95.5 | 97.4 |
| External sample validation (n=127)<sup>a</sup> |  |  |  |  |  |  |  |  |  |
| One-field level, % | 99.3 | 94.5 | 98.3 | 94.1 | 96.9 | 99.3 | 99.3 | 96.9 | 97.1 |
| Two-field level, % | 97.2 | 93.1 | 98.3 | 89.3 | 96.9 | 98.3 | 97.9 | 91.7 | 95.3 |
| G-group level, % | 96.9 | 92.8 | 97.6 | 87.9 | 96.9 | 98.3 | 97.9 | 91.7 | 95.0 |
| C4 |  |  |  |  |  |  |  |  |  |
|  | C4A |  |  | C4B |  | HERV |  | Total |  |
| Leave-one-out cross validation (n=1,537) |  |  |  |  |  |  |  |  |  |
| Haploid level, % | 94.4 |  |  | 94.7 |  | 94.1 |  | 94.4 |  |
| Diploid level, % (r<sup>2</sup>)<sup>b</sup> | 90.1 (0.80) |  |  | 90.1 (0.80) |  | 91.5 (0.90) |  | 90.5 (0.83) |  |
| External sample validation (n=127)<sup>c</sup> |  |  |  |  |  |  |  |  |  |
| Diploid level, % | 90.3 |  |  | 89.7 |  | 90.3 |  | 90.1 |  |

<sup>a</sup>HLA alleles imputed from MHC SNPs were compared with those genotyped from WGS data to calculate concordance rates in 127 external non-reference samples.
<sup>b</sup>Pearson correlation coefficients were calculated between typed (based on WGS data) and imputed C4-related copy numbers using reference samples.
<sup>c</sup>Concordance rates were calculated between ddPCR-typed and imputed copy numbers for C4A, C4B, and HERV.

For *C4*-related CNVs, our imputation also achieved highly reliable concordance rates for haploid copy numbers, showing 94.4% for *C4A*, 94.4% for *C4B*, and 94.1% for HERV, bringing the overall accuracy to 94.4%. As anticipated, the imputed diploid copy numbers were highly consistent with the observed diploid copy number, yielding an overall accuracy of 90.2% (r^2^=0.83) at the diploid level (**Table 1**), which was slightly better than a previous *C4* imputation approach (e.g., r^2^ values of 0.78 and 0.65 for *C4A* in Europeans and Africans, respectively)^18^. Additionally, the vast majority (95.2%) of discordant results showed only one-copy differences between imputed and observed copy numbers (**Supplementary Tables 3** and **4**).

### Application of the MHC reference panel to large-scale SLE GWAS datasets: imputation and accuracy assessment

We implemented the new MHC reference panel to simultaneously impute various MHC variants, including HLA and *C4* variants, across the MHC region on chromosome 6, using SNP genotypes from two Korean SLE GWAS datasets (discovery dataset: 2,023 SLE patients and 71,125 healthy controls; replication dataset: 348 patients and 4,203 controls; **Supplementary Table 5**). The average imputation quality scores, INFO, were reliable, showing 0.92 for HLA alleles with a minor allele frequency over 0.5% in the discovery dataset and 0.90 in the replication dataset. For *C4*-related CNVs, both datasets achieved an average INFO score of 0.81, which surpassed the typical INFO threshold used in GWAS (**Supplementary Fig. 7**).

To assess imputation accuracy, we compared imputed results with directly genotyped results from 127 QC-passed samples, using WGS for HLA classical alleles and ddPCR for *C4*-related CNVs. The concordance rates were high and consistent with those observed in leave-one-out cross-validation of the MHC reference panel (**Table 1**). HLA alleles achieved an average concordance of 95.3% at two-field resolution, and *C4*-related CNVs achieved 89.8% at the diploid copy number level.

Additionally, we investigated the correlation between imputed diploid copy numbers of *C4*-related elements and plasma C4 protein levels in 1,284 patents with SLE, adjusting for covariates including SLE disease activity index (SLEDAI), sex, and the top five genotypic principal components (PCs). Our correlation analysis using imputed CNVs replicated the known effects of *C4*-related CNVs on *C4* expression, as higher plasma C4 protein levels were strongly associated with increased copy numbers of *C4* and decreased copy numbers of HERV inserts (*P*=8.8×10^-69^ for *C4* and *P*=2.9×10^-23^ for HERV; **Fig. 4** and **Supplementary Table 6**). This finding reinforces the reliability of our C4 CNV imputation. Notably, we found no evidence that CNVs of *C4A* and *C4B* have differential regulatory effects on C4 production (**Supplementary Table 6**).

**Fig. 4.**
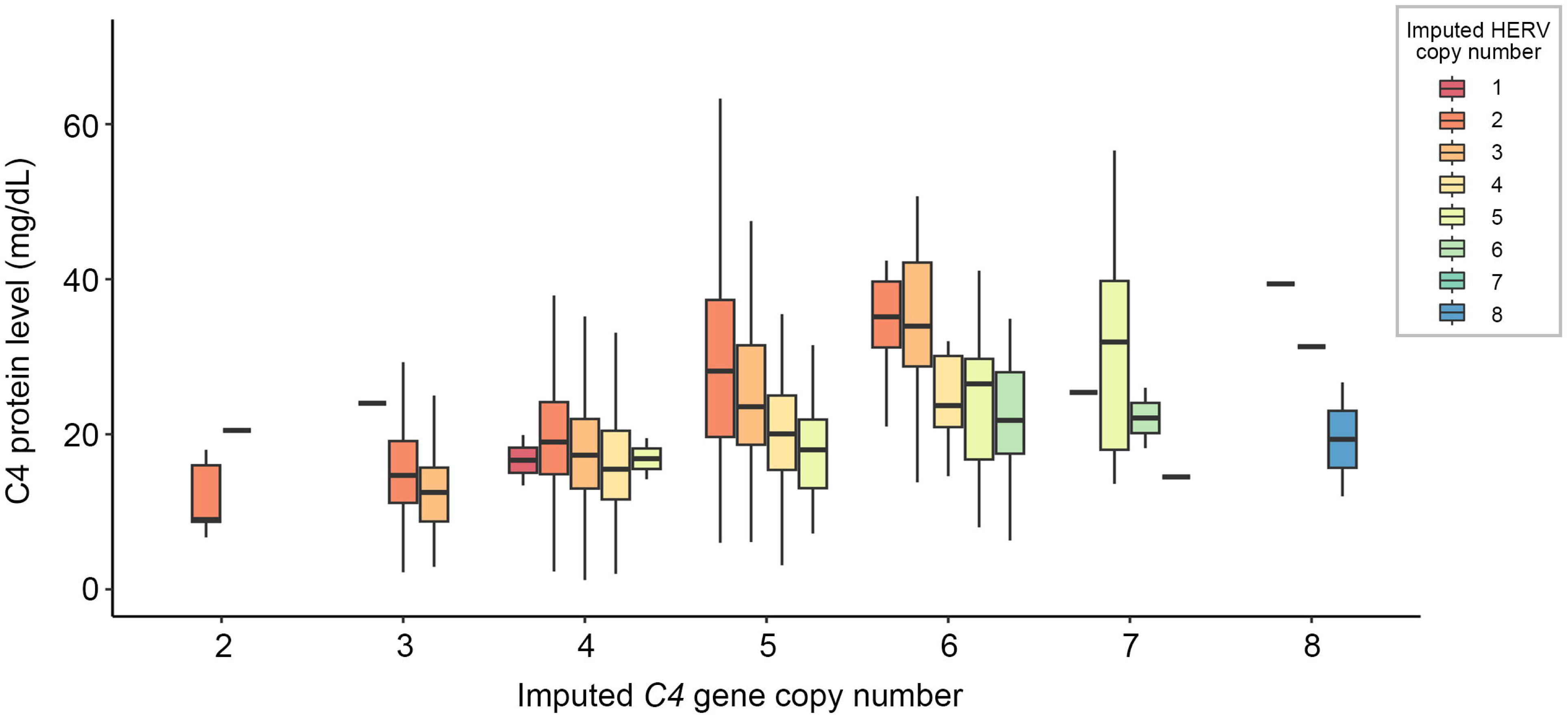
Association of plasma C4 protein levels with imputed *C4* and HERV copy numbers in SLE patients. The boxplot displays the relationship between plasma C4 protein levels and imputed *C4* copy numbers, further stratified by imputed HERV copy numbers in 1,284 SLE patients. A significant positive correlation is observed between *C4* copy numbers and protein levels (*P*=8.8×10^-69^), while HERV copy numbers show a negative correlation (*P*=2.9×10^-23^). One outlier with C4 protein levels over 100mg/dL from a sample with six HERV-inserted *C4* copies was omitted from the plot.

### Primary SLE association at *HLA-DRB1* tagged by five amino acids in the epitope-binding groove

We performed a stepwise conditional regression analysis to unravel multiple independent SLE association signals across the MHC region (28-34Mb on chromosome 6; hg38) using the discovery GWAS dataset. The genetic associations of HLA classical alleles, HLA amino acid residues, SNPs, indels, and the diploid and haploid copy numbers of *C4*-related elements with SLE risk were assessed using logistic regression adjusting for the top five principal components and sex. The multi-allelic effect of each HLA amino acid position was evaluated by a likelihood ratio test (LRT), comparing a full model that included amino acid residues with a nested model that excluded these residues.

We identified the primary association signal at *HLA-DRB1*, with the most significant association occurring at amino acid position 13, which is occupied by six possible residues (*P*=3.16×10^-59^; **Figs. 5a**, **6a** and **6g**). Position 11, also including six possible residues, demonstrated strong LD with position 13 (r^2^=0.81) and showed a highly significant association (*P*=1.58×10^-49^; **Fig. 5a** and **Fig. 6a, g**). Among the HLA classical alleles, *HLA-DRB1*15:01:01G* (*P*=4.90×10^-50^; **Fig 5a**) exhibited the most significant association with SLE risk, followed by *HLA-DQB1*06:02:01G* (*P*=7.11×10^-48^; **Fig 5a**), both of which are well-established as SLE-risk alleles in East Asian populations.

**Fig. 5.**
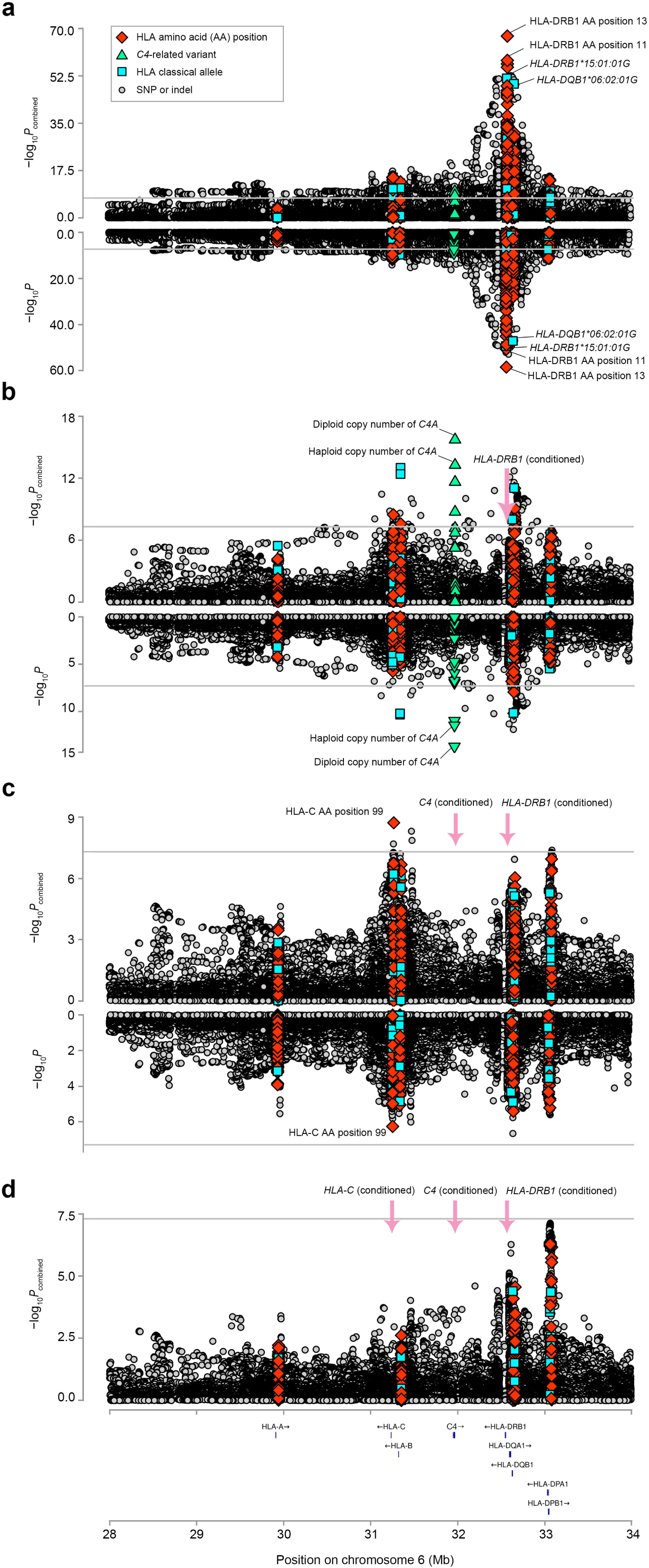
Independent SLE association signals at *HLA-DRB1*, *C4*, and *HLA-C*. MHC-SLE association results from stepwise conditional regression analyses are shown in mirror regional association plots. The lower section of each mirror plot represents analysis results from the discovery GWAS dataset, while the upper section shows results from combined analyses incorporating both discovery and replication GWAS datasets. Significance levels were calculated using logistic regression for binary-coded variables (e,g., presence of an HLA classical allele or a *C4*- related haploid copy number allele) and for diploid copy numbers of *C4*-related elements, and using LRTs for multi-allelic variants (e.g., HLA amino acid positions with multiple residues, *C4*-related CNVs with multiple haploid copy number alleles). The gray horizontal line indicates the genome-wide significance threshold (*P*=5.0×10^-8^). **a.** Unconditional analysis identified the strongest association within *HLA-DRB1*. Amino acid positions 13 exhibited the highest significance (*P*=3.16×10^-59^; *P*_combined_=5.35×10^-68^) followed by position 11 (*P*=1.58×10^-49^; *P*_combined_=1.82×10^-56^), while *HLA- DRB1*15:01:01G* was the most significantly associated allele (*P*=4.90×10^-50^; *P*_combined_=2.51×10^-52^). **b.** After conditioning on all G-group classical alleles of *HLA-DRB1*, a secondary signal was uncovered at *C4*, with the diploid copy number (*P*=1.97×10^-14^; *P*_combined_=1.86×10^-16^) and haploid copy number (*P*=3.34×10^-12^; *P*_combined_=5.34×10^-14^) of the *C4A* allele showing the strongest associations. **c.** Conditioning on all G-group *HLA-DRB1* classical alleles and diploid copy numbers of all *C4* elements (*C4A*, *C4B*, and HERV) revealed a tertiary signal within *HLA-C* in the combined analysis, with amino acid position 99 exhibiting the most significant association (*P*=1.87×10^-9^). No tertiary signal was detected in the discovery GWAS dataset alone. **d.** After conditioning on *HLA-DRB1*, *C4*, and *HLA-C* in the combined analysis, no significant variants were identified.

**Fig. 6.**
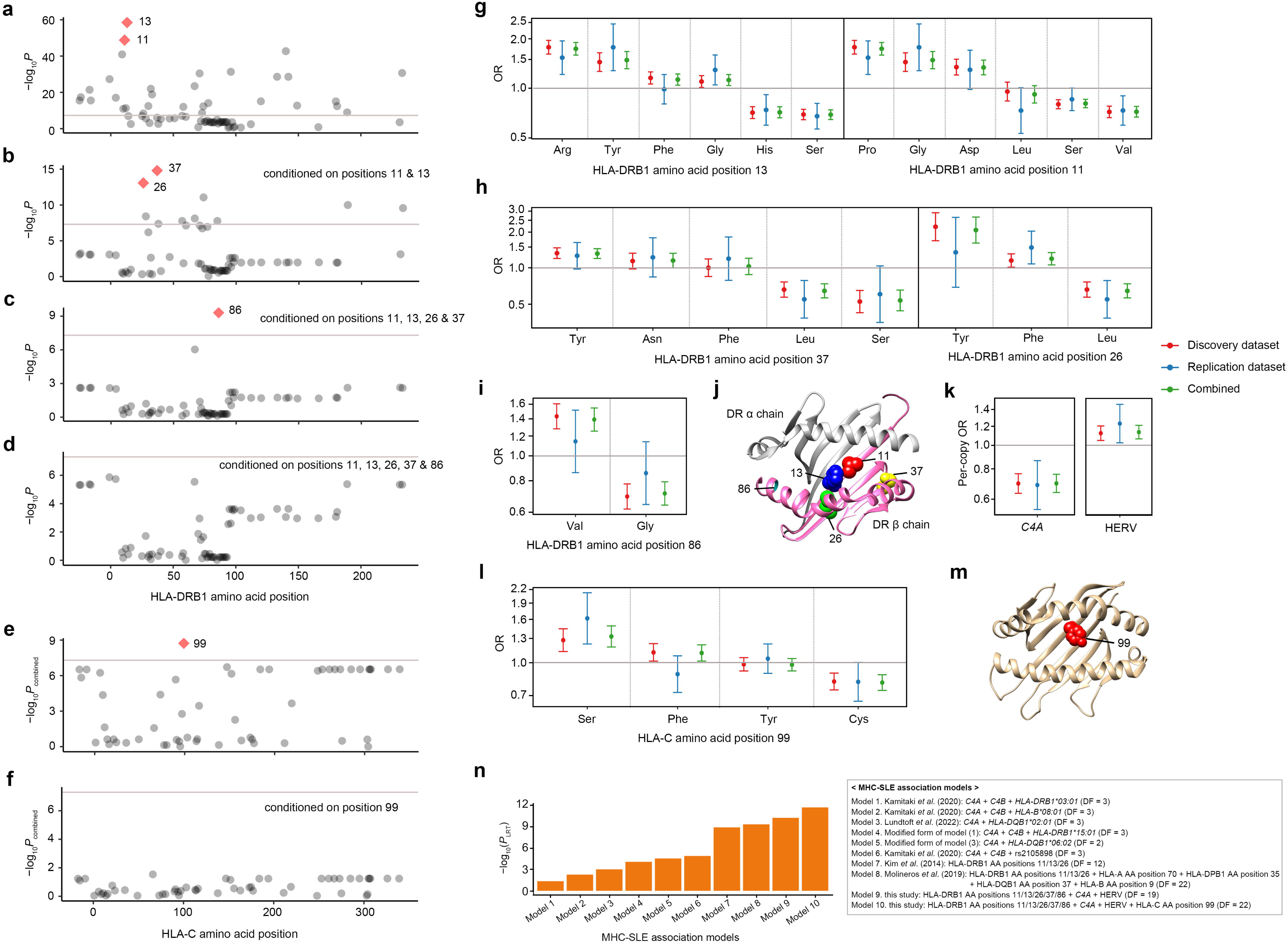
Key SLE-associated variants within *HLA-DRB1*, *C4*, and *HLA-C*. **a-f.** Stepwise conditional analyses at the amino acid level pinpointed independently SLE-associated positions, with HLA-DRB1 positions identified in the discovery dataset and HLA-C positions identified in the combined discovery and replication datasets. **a.** Position 13 and its correlated position 11 were identified in HLA-DRB1. **b.** Conditioning on positions 13 and 11 identified independent signals at position 37 (or 26). **c.** Position 86 was identified when conditioned on positions 13, 11, 37, and 26. **d.** No significant positions remained in HLA-DRB1 after conditioning on all five identified positions (*P*<5.0×10^-8^). **e.** Conditioning on *HLA-DRB1* and *C4* identified HLA-C amino acid position 99. **f.** No additional HLA-C positions were significant after conditioning on position 99 along with *HLA-DRB1* and *C4*. **g-i.** ORs (dots) with 95% CIs (error bars) for amino acid residues, observed in stepwise conditional analysis, are plotted for HLA-DRB1 positions 13 and 11 **(g)**, positions 37 and 26 **(h)**, and position 86 **(i)**, across datasets: discovery (n=73,148, red), replication (n=4,551, blue), and combined (n=77,699, green). **j.** The five identified amino acid positions are located on the epitope-binding groove of the HLA-DR beta chain. **k.** Per-copy ORs with 95% CIs for *C4A* and HERV are plotted across datasets. Effect estimates for *C4A* were adjusted for *HLA-DRB1* and other covariates, while those for HERV were adjusted for *HLA-DRB1*, *C4A*, and the same set of covariates. **l.** ORs with 95% CIs for residues at HLA-C position 99, conditioned on *HLA-DRB1* and *C4*, are plotted across datasets. **m.** Amino acid position 99 is located on the epitope-binding groove of HLA-C. **n.** *P*_LRT_ values from the replication dataset for previously proposed and our MHC-SLE association models are plotted. To address potential overfitting, Model 9 was derived from the discovery dataset, excluding the *HLA-C* signal. Model 10 represents the full association model of this study, including the *HLA-C* signal. Our MHC-SLE association models, containing the identified signals, achieved the highest significance whether or not the *HLA-C* signal was included (*P*_Model_9_=5.50×10^-11^; *P*_Model_10_=1.91×10^-12^). AA, amino acid; DF, degree of freedom.

Given the complex nature of long-range haplotypes, which may include key amino acid combinations involving residues at positions 13 and 11 that tag specific *HLA-DRB1* classical alleles and drive the SLE association of *HLA-DRB1*, we further investigated additional independent associations within *HLA-DRB1* at the amino acid level. After accounting for the effects of the tightly correlated amino acid positions 13 and 11, we identified significant associations at amino acid positions 37 (*P*=1.54×10^-15^) and 26 (*P*=8.36×10^-14^), which showed moderate residue correlation (r^2^=0.42) (**Fig. 6b, h**). These positions provided similar model fits for SLE association (*P*_LRT_=1.59×10^-71^ for positions 11-13-37 and *P*_LRT_=8.65×10^-70^ for positions 11-13-26), which has already been reported in previous studies^5,6^. Considering the uncertainty of the secondary SLE-risk amino acid position, we accounted for both positions 26 and 37 in the next round of stepwise conditional regression. After conditioning on the effects of positions 11, 13, 26, and 37, we newly identified amino acid position 86 (*P*=4.93×10^-10^; **Fig. 6c, i**). Adding or excluding position 26 in the conditional regression model did not change the statistical significance of position 86. No additional significant positions were detected after conditioning on the amino acid position 11, 13, 26, 37, and 86 (**Fig. 6d**).

The associations at these positions were further examined in the replication GWAS dataset, revealing highly consistent residue-specific effect sizes across both the discovery and replication GWAS datasets (**Fig. 6g-i**) and increased significance in the combined analysis of both datasets (**Supplementary Data 1**). Moreover, the three signals derived from the five identified amino acid positions exhibited the best SLE association model fit among all possible combinations of HLA-DRB1 amino acid positions in LRTs, accounting for the differences in the number of residues as indicated by the degree of freedom (**Supplementary Fig. 8**). These positions are located in close spatial proximity within the epitope-binding pocket of HLA-DR molecules (**Fig. 6j**).

Based on these findings, amino acid haplotypes of five key positions were tested for SLE associations (**Table 2**). Sixteen haplotypes with frequency > 0.5% were identified, including five SLE-risk and seven SLE-protective haplotypes (*P*_combined_ < 0.05). The amino acid haplotype model effectively explained associations of various classical alleles, such as *HLA-DRB1*15:01* with the PRSFV haplotype. Furthermore, *HLA-DRB1*03:01* (SSYNV) and *HLA-DRB1*15:03* (PRSFV), known to be specific to Europeans and Africans, respectively, were identified as having SLE-risk amino acid haplotypes, even though these classical alleles were present at low or absent frequencies in in the Korean population. Similarly, *HLA-DRB1*13:02* and *HLA-DRB1*14:03*, previously reported as significant SLE-protective alleles^28^, were found to have the SLE-protective haplotype SSPNG. Taken together, this suggests that amino acid haplotypes not only provide a parsimonious disease association model compared to the conventional model using classical alleles but also demonstrate a highly transferable model across different ancestries.

**Table 2.** Association of HLA amino acid haplotypes with susceptibility to SLE.

| HLA-DRB1 amino acid position |  |  |  |  | Frequency |  | SLE association |  | Observed classical *HLA-DRB1* alleles |
| --- | --- | --- | --- | --- | --- | --- | --- | --- | --- |
| 11 | 13 | 26 | 37 | 86 | Cases | Controls | OR (95% CI) | *P*<sub>combined</sub> |  |
| Pro | Arg | Phe | Ser | Val | 0.145 | 0.084 | 1.85 (1.70 - 2.01) | 6.06×10<sup>-46</sup> | **\*15:01**, *15:13 |
| Gly | Tyr | Phe | Phe | Gly | 0.089 | 0.066 | 1.39 (1.26 - 1.54) | 3.81×10<sup>-10</sup> | *07:01 |
| Ser | Gly | Phe | Tyr | Gly | 0.129 | 0.100 | 1.34 (1.22 - 1.46) | 8.55×10<sup>-11</sup> | *08:01, *08:02, *08:03 |
| Asp | Phe | Tyr | Asn | Gly | 0.119 | 0.095 | 1.29 (1.18 - 1.42) | 2.47×10<sup>-8</sup> | *09:01, *09:05 |
| Ser | Ser | Tyr | Asn | Val | 0.024 | 0.019 | 1.25 (1.03 - 1.51) | 2.24×10<sup>-2</sup> | **\*03:01** |
| Pro | Arg | Phe | Ser | Gly | 0.043 | 0.044 | 0.99 (0.86 - 1.15) | 9.37×10<sup>-1</sup> | *15:02, *16:02, *16:09 |
| Leu | Phe | Leu | Ser | Gly | 0.057 | 0.062 | 0.92 (0.81 - 1.04) | 1.74×10<sup>-1</sup> | *01:01 |
| Val | Phe | Leu | Tyr | Gly | 0.014 | 0.016 | 0.87 (0.68 - 1.11) | 2.56×10<sup>-1</sup> | *10:01 |
| Ser | Ser | Phe | Asn | Val | 0.022 | 0.025 | 0.86 (0.71 - 1.05) | 1.49×10<sup>-1</sup> | *13:01, *14:06, *14:12 |
| Val | His | Phe | Tyr | Gly | 0.077 | 0.091 | 0.84 (0.75 - 0.93) | 1.38×10<sup>-3</sup> | *04:01, *04:05, *04:07, *04:08 |
| Ser | Gly | Leu | Leu | Val | 0.063 | 0.077 | 0.81 (0.72 - 0.91) | 4.03×10<sup>-4</sup> | *12:01, *12:02 |
| Val | His | Phe | Tyr | Val | 0.037 | 0.047 | 0.79 (0.68 - 0.92) | 2.31×10<sup>-3</sup> | *04:03, *04:04, *04:10 |
| Ser | Ser | Phe | Phe | Val | 0.046 | 0.058 | 0.77 (0.67 - 0.88) | 2.08×10<sup>-4</sup> | *14:01, *14:05 |
| Ser | Ser | Phe | Asn | Gly | 0.066 | 0.102 | 0.61 (0.55 - 0.69) | 2.37×10<sup>-16</sup> | *13:02, *13:39, *14:02, *14:03 |
| Ser | Ser | Phe | Tyr | Gly | 0.027 | 0.046 | 0.57 (0.48 - 0.68) | 6.01×10<sup>-10</sup> | *11:01, *11:11, *11:19, *11:39, *13:07 |
| Val | His | Phe | Ser | Val | 0.026 | 0.052 | 0.49 (0.41 - 0.58) | 9.83×10<sup>-15</sup> | *04:06 |
| HLA-C amino acid position 99 |  |  |  |  |  |  |  |  | Observed classical *HLA-C* alleles |
|  |  | Ser |  |  | 0.108 | 0.084 | 1.29 (1.16 - 1.44) | 2.24×10<sup>-6</sup> | *07:02 |
|  |  | Phe |  |  | 0.171 | 0.193 | 1.10 (1.01 - 1.20) | 2.56×10<sup>-2</sup> | *04:01, *04:03, *14:02, *14:03 |
|  |  | Tyr |  |  | 0.561 | 0.544 | 0.97 (0.90 - 1.03) | 3.02×10<sup>-1</sup> | *02:02, *03:02, *03:03, *03:04, *03:132, *03:24, *03:43, *03:49, *05:01, *06:02, *06:12, *07:01, *07:04, *07:151, *08:01, *08:02, *08:03, *08:06, *08:10, *12:02, *12:03, *12:05, *15:02, *15:05, *15:62 |
|  |  | Cys |  |  | 0.154 | 0.176 | 0.79 (0.72 - 0.86) | 1.31×10<sup>-7</sup> | *01:02, *01:03, *15:102 |

HLA amino acid haplotypes with frequencies over 0.5% are listed. Haplotype-specific odds ratios (OR) of having SLE were calculated for each HLA-DRB1 amino acid haplotype by logistic regression adjusting for dataset, sex, and the top five genotype principal components. Two well-validated SLE-risk *HLA-DRB1* alleles are highlighted in bold. Residue-specific ORs of HLA-C amino acid position 99 were calculated by logistic regression, conditioning on G-group *HLA-DRB1* alleles and the diploid copy number of *C4A*, *C4B*, and *HERV*, while adjusting for dataset, sex, and the top five genotype principal components.
SLE, systemic lupus erythematosus; OR, odds ratio; CI, confidence interval.

### Dissecting the secondary SLE association revealed that decreased *C4A* and increased HERV copy numbers confer the risk of SLE, independent of *HLA-DRB1*

To determine if there were additional independent MHC associations beyond *HLA-DRB1*, we conducted a conditional analysis that adjusted for all *HLA-DRB1* G-group alleles with frequencies greater than 0.5%, thereby eliminating the influence of *HLA-DRB1* on the SLE associations. Using the discovery GWAS dataset, we identified a secondary signal within the *C4* and observed moderate correlation with *HLA-DRB1*, which was adequately controlled for in the logistic regression model (**Fig. 5b** and **Supplementary Table 7**). We found that the diploid copy numbers of *C4A* showed the most significant association in the conditional analysis (*P*=1.97×10^-14^), which was much stronger than the copy numbers of *C4B* (*P*=1.09×10^-7^) and *HERV* (*P*=1.85×10^-2^), as well as the total gene copy number of *C4* (*P*=5.37×10^-2^) (**Fig. 5b**). The lower copy number of *C4A* was associated with a higher risk of SLE, with a per-copy odds ratio (OR) of 0.70 with a 95% confidence interval (CI) from 0.63 to 0.76 (**Fig. 6k**). This association pattern was consistently observed at the haploid-level, with *C4A* showing the strongest significance among the *C4*-related CNVs (*P*=3.34×10^-12^; **Fig. 5b**). Compared with one haploid copy of *C4A* as the reference, the deficiency of *C4A* increased SLE risk (OR=1.36, 95% CI=1.18—1.57, *P*=2.38×10^-5^), while the gain of *C4A* copy showed a protective effect (OR=0.75, 95% CI=0.67—0.83, *P*=8.87×10^-8^).

As previously known, the haploid-level copy numbers of *C4A*, *C4B* and HERV are in LD with each other (**Supplementary Table 1**). To understand the overall effect of *C4*-related CNVs and estimate the mutually independent SLE-risk effects of each *C4*-related CNV, we conducted a joint conditional regression analysis using the diploid copy number of *C4A*, *C4B*, and HERV, adjusting for *HLA-DRB1* and genetic background (**Table 3**). This joint model revealed a significant contribution of *C4*-related CNVs to SLE susceptibility (*P*_LRT_=1.80×10^-15^), exhibiting significant, independent associations of *C4A* (*P*=4.12×10^-11^, per-copy OR=0.69, 95% CI= 0.62 –0.77) and HERV (*P*=1.13×10^-2^, per-copy OR=1.10, 95% CI= 1.02 –1.19), but not *C4B* (*P*=0.24). In contrast to the effects of the *C4A* CNV, the copy number of HERV was positively associated with the risk of SLE (**Fig. 6k**). However, the known biological effects of both decreased *C4A* copy number and increased HERV copy number are the same, collectively resulting in decreased *C4A* expression, as supported by our correlation analysis between plasma C4 protein and the copy number of *C4* and HERV in our study cohort (**Fig. 4c**).

**Table 3.**
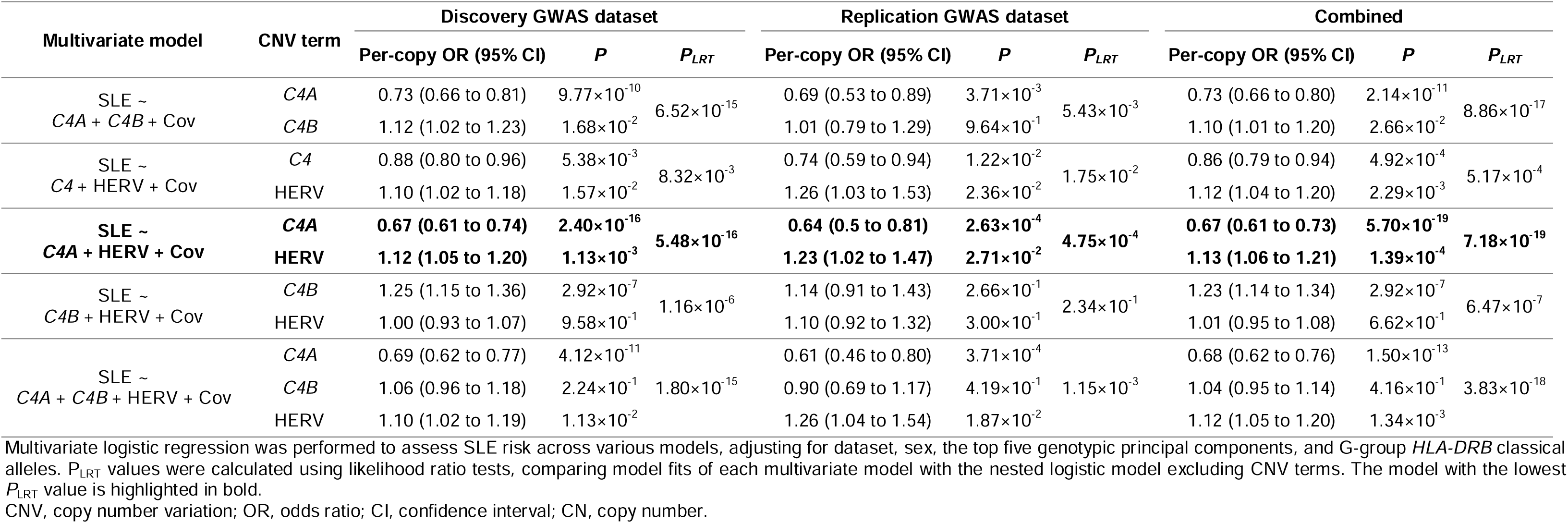
Joint effects of *C4*-related CNVs on SLE risk, adjusted for *HLA-DRB1* effects.

All *C4*-related association signals involving *C4A* and HERV were robustly validated in the replication GWAS dataset, and demonstrated increased significance in combined analyses incorporating both discovery and replication GWAS datasets (**Fig. 5b**, **6k**, **Table 3**, and **Supplementary Data 1**).

We also investigated previously reported sex bias in *C4*-SLE associations using a logistic regression model with an interaction term between sex and *C4*-related CNVs, but found no evidence of sex-biased effect sizes on SLE (**Supplementary Table 8**). Indeed, plasma C4 protein levels normalized by the *C4* copy number were consistent between males and females in 1,284 SLE patients (**Supplementary Fig. 9** and **Supplementary Table 6**).

### Tertiary association signals at *HLA-C* amino acid position 99

After conditioning on *HLA-DRB1* and *C4*, the tertiary signal was identified within *HLA-C* in the combined analysis but not in the discovery dataset alone, with the most significant association at amino acid position 99 (*P*=5.39 ×10^-7^; *P*_combined_=1.87×10^-9^; **Figs. 5c**, **6e**, **6l** and **Supplementary Data 1**). Position 99 is highly polymorphic, with four possible residues, and its effect sizes were consistent across the discovery and replication datasets (**Fig. 6l**). Located in the center of the peptide-binding groove of HLA-C (**Fig. 6m**), this position has been associated with the risk of other immune-mediated disorders^29,30^. Among the four residues at position 99, serine was associated with an increased risk of SLE, while cysteine was protective (**Table 2**). Serine at position 99 primarily tags *HLA-C*07:02* in Koreans, which belongs to a previously reported long-range SLE-risk haplotype in Europeans, though its independent effect has not been investigated^20,31,32^. Cysteine at position 99 is mostly found in *HLA- C*01:02*, which has been associated with other diseases but not with SLE^33–35^.

### SLE-MHC association model comparison in the Korean population

We then focused on evaluating the fit of our SLE-MHC association model to assess its performance relative to previously proposed SLE-MHC association models^5,6,18,19^. To avoid overfitting, our association model included only the variants identified from the discovery dataset, comprising amino acid positions 11, 13, 26, 37, and 86 in HLA-DRB1, along with the diploid copy numbers of *C4A* and HERV, while excluding HLA-C amino acid position 99 identified in a combined analysis. The model fit was subsequently evaluated in the replication GWAS dataset.

Our model demonstrated a strong association with a *P*_LRT_ value of 5.50×10^-11^ in the replication dataset (**Fig. 6n**), outperforming all previous SLE-MHC association models, as indicated by P_LRT_ values that accounted for differences in the number of variables (or degrees of freedom). Notably, some previous models included European-specific alleles, such as *HLA-DRB1*03:01* or *HLA- DQB1*02:01*. To account for this, we re-evaluated these models by substituting the European-specific alleles with the East Asian-specific SLE-risk allele *HLA-DRB1*15:01* or its strong proxy allele *HLA-DQB1*06:02* (r^2^=0.94 in Koreans). Although this substitution improved model performance, the association significance level remained weaker than that of our comprehensive MHC model. These findings emphasize the importance of considering the collective effects of multiple *HLA-DRB1* classical alleles (tagged by missense variants) as well as the diploid copy numbers of *C4A* and HERV. Additionally, when we incorporated the tertiary association at amino acid position 99 of HLA-C, identified only through the combined datasets, the fit of our association model improved, as expected (*P*=1.91×10^-12^; **Fig. 6n**).

## Discussion

This study delved into the genetic associations within the MHC region using two independent Korean SLE case-control datasets, leveraging a newly developed MHC imputation reference panel. Our findings underscore the significant, independent associations of *HLA-DRB1*, *C4*, and *HLA-C* with the risk of SLE. The MHC-SLE association model we proposed outperformed other models comprising variants reported in previous MHC fine-mapping studies. Model comparisons were conducted on an independent replication dataset based on LRTs that accounted for differences in degrees of freedom, ensuring fair and robust comparisons.

The associations of *HLA-DRB1* and *HLA-C* were predominantly observed at specific amino acid positions rather than HLA classical alleles, emphasizing the disease relevance of the missense variants that shape the surface of epitope-binding grooves of *HLA-DRB1* and *HLA-C* in SLE. Amino acid positions 11, 13, 26, and 37 of HLA-DRB1 have been consistently highlighted in previous HLA fine-mapping studies^5,6^, and we have now identified an additional position, 86, which collectively accounts for most of the genetic contribution of *HLA-DRB1*. Position 86 has been linked to the risk of type 1 diabetes^36^. These five amino acid positions of HLA-DRB1, representing three independent association signals, provided the simplest and ancestrally transferable explanation, supporting the associations of various classical alleles. Notably, the associations of HLA-DRB1 amino acid haplotypes well align with the ancestry-specific associations of *HLA-DRB1* (e.g., *HLA-DRB1*03:01* in Europeans and *HLA-DRB1*15:03* in Africans).

Importantly, this is the first study to investigate the disease effects of *C4* CNVs while adjusting for the multiallelic effects of *HLA-DRB1*. There has been debate regarding whether *HLA-DRB1* or *C4* is the primary driver of SLE in the MHC region, and whether the associations of SLE with *HLA-DRB1* and *C4*, which are in LD, are independent of each other^37^. Our conditional association model, which controlled for the influence of *HLA-DRB1* on SLE risk, identified the most significant association at *C4*, revealing that a lower copy number of *C4A* and, for the first time, a higher copy number of HERV were independently associated with an increased risk of SLE. Plasma C4 analysis indicated that C4 proteins levels are predominantly influenced by the total copy number of *C4*, with no evidence of allele-specific regulatory effects between *C4A* and *C4B*. Additionally, a HERV element, inserted within *C4*, leads to down-regulation of plasma C4 proteins. Considering both the genetic associations with SLE and protein levels, these findings suggest that the reduced production of the C4A protein isoform, functionally distinct from the C4B isoform, is driven by both low *C4A* copy numbers and HERV- mediated attenuation. This reduction plays a crucial role in SLE pathogenesis, particularly through *C4A*-specific pathways such as the regulation of autoreactive B cells. A recent study using a mouse model showed that the C4A isoform is more efficient than C4B at binding and removing apoptotic debris, enhancing apoptotic clearance, and reducing autoantigen exposure, which limits the activation of autoreactive B cells^38^. It also found that C4A regulates germinal centers by restricting the expansion of these B cells^38^. These findings highlight the role of *C4A* for maintaining immune tolerance and preventing pathogenic autoimmunity in SLE.

Our fine-mapping study was particularly successful by leveraging the relatively less extensive LD in the Korean population compared to Europeans, along with the use of a high-accuracy MHC imputation reference panel. Previous studies have demonstrated that European populations exhibits larger and more extended LD within the MHC region compared to African and Asian populations, likely due to population-specific factors such as lower recombination rates, resulting in slower decay of LD in Europeans^18,19,39–42^. Consistently, European SLE-MHC association analyses have reported long-range SLE-risk haplotypes involving multiple HLA genes in extensive LD, spanning MHC class I and II regions, such as the *C*07:01*–*B*08:01*–*DRB1*03:01*–*DQB1*02:01* haplotype^19,31^. In contrast, Asian and African populations have shown less extensive LD patterns, with relatively more predominant SLE associations in the MHC class II region^20,21^.

Indeed, recent analyses by Kamitaki *et al.*, whose study is detailed in the Introduction section, found that the significance of *C4* CNV effects was comparable to that of well-known risk classical alleles in Europeans^18,19^. However, the strong correlations between *C4* and HLA in Europeans posed challenges in distinguishing the independent effect of *C4*, especially considering the strong correlation between the SLE-risk *C4* haplotype (single-copy, short-form *C4B*) and SLE-risk HLA alleles, including *HLA-DRB1*03:01*, *HLA-DRB1*08:01*, and *HLA-DQB1*02:01* (r^2^>0.7)^18,19,39^. Conversely, findings from African populations revealed weaker correlations between *C4* and HLA variants (maximum r^2^ between *C4* haplotypes and HLA alleles = 0.31), making it easier to pinpoint the independent association signal of *C4*. However, in their study, the imputation accuracy for *C4* in Africans was quite poor, and their conditional model controlling only for *HLA-DRB1*03:01* does not fully support their conclusion that the primary signal driving the risk of SLE is derived from *C4*^18^.

In Asian populations, the particularly strong association between *HLA-DRB1* and SLE risk has been well-established^5,6^. In our current study, the association of *C4* was initially overshadowed by the predominant effect of *HLA-DRB1* in the unconditional analysis (*P*_combined_ of *C4A*=1.48×10^-6^). However, correlations between *HLA-DRB1* classical alleles and *C4*-related haploid copy numbers were moderated (maximum r^2^ between *C4*-related haploid copy numbers and HLA alleles = 0.39; **Supplementary Table 7**), our conditional analysis enabled us to pinpoint the secondary association signal at *C4*.

We developed the first MHC imputation reference panel capable of simultaneously imputing various HLA variants, *C4*-related CNVs, and other MHC variants. The *C4* gene, despite its biological relevance to autoimmune diseases like SLE, has been relatively underexplored compared to HLA genes, largely due to technically complex and low-throughput methods required to characterize its genetic structure. In the present study, we leveraged WGS data from large-scale general Korean cohorts to characterize *C4* alleles and CNVs using an existing reliable computational approach^18,23^.

One of the major challenges was accurately defining haploid-level copy numbers from WGS-derived diploid copy numbers. To address this, we employed haplotype clustering based on SNPs and short indels around the *C4* gene, utilizing deep learning-based models and an iterative optimization method to determine the most likely copy numbers of *C4A*, *C4B*, and HERV for each haplotype cluster. This method not only allowed us to construct an MHC imputation reference panel with high accuracy for C4-related CNVs, but it also holds significant potential for broader applications in other studies dealing with complex CNV genotyping. Nonetheless, Our imputation panel has a limitation that it only includes copy number information for *C4A*, *C4B*, and HERV in each homologous chromosome pair, leading to ambiguity in determining how many copies of *C4A* and *C4B* are in the long form with the HERV insert. This ambiguity is particularly pronounced in haplotypes where both *C4A* and *C4B* alleles are present but the number of HERV copies is fewer than the combined total of *C4A* and *C4B*.

To validate the imputation performance of our reference panel, we rigorously evaluated the imputation accuracy through various methods, including cross-validation with internal and external samples, that compares imputed data with actual data from WGS and ddPCR technologies. These evaluations consistently demonstrated superior imputation accuracy for both HLA variants and *C4*- related CNVs. We have made this imputation panel publicly available via an imputation server (https://coda.nih.go.kr/usab/kis/intro.do), providing other researchers with a valuable tool to better understand causal variants in the MHC region, especially for inflammatory diseases in East Asian populations.

In summary, we have demonstrated an enhanced SLE-MHC association model from the Korean population through the development of a novel MHC imputation reference panel. This advancement not only enriches our understanding of the biological mechanisms underlying the role of HLA and *C4* in SLE pathogenesis but also holds promise for refining genetic risk assessments for SLE. Furthermore, the application of our MHC panel is poised to significantly advance the investigation of MHC associations in other immune-mediated disorders within East Asian populations, paving the way for further insights and progress in personalized medicine.

## Methods

### Cohorts for constructing the MHC imputation reference panel and fine-mapping MHC-SLE associations

Two independent WGS batches from 1,904 unrelated healthy Korean individuals were utilized for constructing the MHC imputation reference panel. These batches were generated by the Ulsan National Institute of Science and Technology (UNIST) (reference batch #1; n=904; published data^43^) and the Korea Reference Genome (KRG) project, led by the National Institute of Health, Republic of Korea (KNIH) (reference batch #2; n=1,000; unpublished data).

The KRG project, which aimed at advancing Korean genome research and identifying a wide spectrum of variants in the Korean population^44^, generated WGS data from around 8,000 individuals. From this dataset, sequencing data from 1,000 randomly selected participants was used to construct the MHC imputation reference panel. All participants provided written informed consent. Genomic DNA samples from the participants was sequenced using the Illumina NovaSeq platform (average depth ∼30x). Raw sequencing data was trimmed using the Trimmomatic software (version 0.39)^45^.

All analyses using the WGS data were conducted under the regulations approved by Institutional Review Board (IRBs) of the UNIST (UNISTIRB-15-19-A, UNISTIRB-16-13-C) and the Korean Disease Control and Prevention Agency (2022-02-04-P-A, KDCA-2024-02-09-R-01).

To characterize MHC associations in SLE, two Korean SLE case-control datasets (discovery and replication GWAS datasets) were used (**Supplementary Table 5**). These datasets were expanded from our recent GWAS datasets^46^ by adding 146 cases 38,305 controls to the previous discovery dataset. Patients with SLE were recruited from Hanyang University Hospital for Rheumatic Diseases and fulfilled either the revised American College of Rheumatology classification criteria or the Systemic Lupus International Collaborating Clinics classification criteria for SLE^47,48^. Healthy controls were recruited from the Korea National Institute of Health. All participants provided written informed consent, and the study protocol was approved by the IRBs at participating institutions (HY-12-005-16, HY-16-07-20, HY-16-129-15).

### Calling SNPs and indels in the MHC region using WGS data

Illumina high-coverage whole-genome 101 bp paired-end sequencing data, with a mean read depth of 29.0 in reference batch #1 and 37.5 in batch #2, were aligned to the human genome reference assembly (GRCh38) using BWA^49^. SNP and indels were called using the Genome Analysis Toolkit (GATK) pipeline^50^. The variants called from the WGS data were filtered for each dataset to retain biallelic variants that met the following QC thresholds: minimum depth of 15 (--minDP) and minimum genotype quality of 20 (--minGQ), a variant quality score at least 20 (--minQ), minor allele frequency over 0.5%, call rate over 90%, and conformity to HWE (*P* > 1×10^-7^). After filtering, a total of 52,675 QC’ed MHC variants (25-35 Mb on chromosome 6; hg38) were retained across both datasets. To further ensure data reliability, we removed variants that showed a batch effect in genotype frequency distribution between the two datasets using Fisher’s exact test (*P* < 0.001), leaving 52,602 variants for further analysis.

### HLA typing at G-group resolution using WGS data

We determined the classical alleles of eight major HLA genes (*HLA-A, -B, -C, -DPA1, -DPB1, -DQA1, -DQB1, -DRB1*) at the G-group resolution using HLA-LA^22^. To ensure high-quality data in the MHC imputation reference panel, we retained reference samples that had an average coverage of over 15× for each gene, resulting in 846 samples from reference batch #1 and 1,000 samples from batch #2 (**Supplementary Fig. 1**). We excluded 12 samples with *HLA-DPA1*01:11*, which were found exclusively in batch #1. We converted these G-group HLA classical alleles into one- and two-field HLA alleles using the NomenCleaner module in HLA-TAPAS^51^. Additionally, HLA amino acid residues and exonic alleles were inferred based on the two-field HLA classical alleles.

### Quantifying diploid copy numbers of *C4*, *C4A*, *C4B*, and HERV using WGS data

We utilized a previously developed method^18^ implemented in Genome STRiP^23^ to estimate the diploid copy numbers of *C4*, *C4A*, *C4B* and HERV by analyzing the read depth across their genic regions and the relative ratio of allele-specific read counts for *C4A* and *C4B*. To ensure that the MHC imputation reference include only highly reliable CNV data, we filtered out samples with a Phred-scale quality score below 20 for any of copy number calls of *C4*-related elements (**Supplementary Fig. 2**). As a result, 1,687 reference samples were retained for the construction of the reference panel, comprising 759 samples from reference batch #1 and 928 samples from batch #2 (**Supplementary Fig. 3**).

### Determination of copy number alleles of *C4A*, *C4B*, and HERV from diploid copy numbers

To accurately determine the CNV genotype (an allelic pair of haploid copy numbers of *C4*-related CNVs) from a given diploid copy number, we first surveyed possible haploid-level copy numbers for *C4A*, *C4B*, and HERV. These were inferred from observed haploid copy numbers in potential genotypes that met a genotype likelihood threshold over 0.01. The genotype likelihood was estimated using the ‘GenerateHaploidCNVGenotypes’ utility in Genome STRiP^23^, which applies a frequency-based expectation-maximization algorithm to estimate the likelihood of a pair of haploid copy numbers under HWE. This analysis showed that the haploid copy numbers could range from 0 to 3 for *C4A* and *C4B*, and from 0 to 4 for HERV.

We then carried out haplotype clustering by examining haplotypes of SNPs and indels around the *C4* region using the HaploNet framework^26^, which uses an unsupervised neural network approach to compress haplotype data into a lower-dimensional space and indirectly cluster haplotypes with a GMVAE model. Input haplotypes were phased with Eagle v2.4.1^52^ and included 601 biallelic variants in the *C4* region (31.7-32.2 Mb on chromosome 6; hg38) pruned at an r^2^ threshold of 0.8 using PLINK^53^. GMVAE models were configured with varying combinations of three parameters: the number of clusters (ranging from 2 to 100), the number of hidden layer nodes (64, 128, 256, 300, and 512), and the number of latent variables (16, 32, 64, and 128; always less than the given number of hidden layer nodes). Each model generated the likelihoods for each haplotype belonging to each identified cluster. For each model, we assigned the haplotypes to their most likely cluster based on the estimated cluster likelihood. Clusters that did not exhibit maximum likelihood for any haplotype were excluded from subsequent analysis steps.

Next, we employed the cyclic coordinate descent method to assign linked haploid copy number of *C4*-related CNVs to each cluster. We initialized a cluster-specific copy number vector, 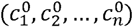, with the most common haploid copy numbers (one for *C4A* and *C4B*, and either one or two for HERV) for all *n* clusters. We iteratively updated these cluster-specific copy numbers by cycling through them one at a time, minimizing a multivariable loss function *f*(*c*) with respect to each cluster-specific copy number while holding the other variables constant. Thus, the copy number of *i*-th cluster at the (*k* + 1)-th iteration 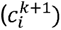 was determined by solving the single variable optimization problem, as follows:

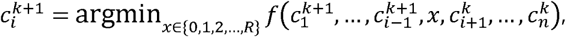

where *f*(*c*) is the 0-1 loss function that measures how many times the predicted diploid copy numbers (=sums of inferred haploid copy numbers per individual) do not match the observed diploid copy number across reference individuals. The iterative optimization continued to update each element of *c* until convergence was achieved, ensuring that the cluster-specific copy numbers best fit the observed diploid copy number data. This analysis was performed separately for *C4A*, *C4B*, and HERV across all GMVAE models, and the model that yielded the best performance was configured with 48 clusters, 64 latent variables, and 300 hidden layer nodes (**Supplementary Fig. 6**). Based on the cluster-wise likelihoods estimated by this model for each haplotype, we determined the CNV genotype of *C4A*, *C4B*, and HERV for each reference individual as the most likely pair of haploid copy numbers that matched the WGS-based diploid copy numbers, and used these CNV genotypes in subsequent analyses. A schematic of the genotyping process is illustrated in **Fig. 3a**.

### Construction of a Korean MHC imputation reference panel

We integrated a wide range of genetic data, including haploid copy numbers of *C4A*, *C4B*, and HERV; HLA classical alleles at one-field, two-field, and G-group resolution; HLA amino acid residues; and other SNPs and indels in the MHC region to assemble an MHC imputation reference panel. All genetic information was encoded in a binary format. Biallelic variants, such as SNPs, indels, and biallelic amino acid positions, were directly represented by their allele names. For multiallelic variants, including HLA classical alleles, multiallelic amino acid positions, and *C4*-related CNVs, we used a binary encoding scheme where each allele or haploid copy number was indicated as its absence (A) or presence (T). For instance, the *C4A* CNV, with possible haploid copy numbers of 0, 1, 2, and 3, was represented by four distinct copy number variables named as C4A_copy0, C4A_copy1, C4A_copy2, and C4A_copy3, respectively. If a sample had 2 and 3 haploid copies, these four variables were encoded as *A*/*A*, *A*/*A*, *A*/*T* (or *T/A*), and *A*/*T* (or *T/A*), respectively. The merged genetic information was phased into haplotypes using Eagle v.2.4.1^52^, including 55,632 biallelic variables in 1,537 samples.

### Evaluation of imputation accuracy of the MHC imputation reference panel

To evaluate the imputation accuracy of our MHC imputation reference panel, we employed a leave-one-out cross-validation strategy using reference samples. For each reference sample, HLA classical alleles and *C4*-related CNVs were imputed based on the genetic data of SNPs and indels using IMPUTE5^54^, with a reference panel composed of all other reference samples. Concordance rates between imputed and genotyped data across all samples were calculated to assess imputation performance.

Additionally, we validated the imputation accuracy using external non-reference samples, randomly selected from the discovery GWAS dataset. High-coverage WGS data were generated for 144 samples to characterize HLA variants and diploid copy numbers of *C4*-related CNVs, using the same analysis and QC methods as for the reference samples. After applying the same filtering criteria, 127 samples with highly reliable calls for HLA and *C4* variants remained for subsequent evaluation of our MHC imputation reference panel (**Supplementary Fig. 1** and **Supplementary Fig. 4**). The diploid copy number of *C4*-related elements were further examined using ddPCR (**Supplementary Fig. 5**), as previously described^24^. *C4* and HLA variants were imputed based on GWAS-array SNPs listed in the discovery GWAS dataset using IMPUTE5, and these imputed results were compared with those experimentally confirmed by ddPCR (for *C4*) and WGS (for HLA). Additionally, the accuracy of WGS-based Genome STRiP-estimated diploid copy numbers of *C4*-related elements was assessed by comparison with ddPCR results (**Fig. 2b**).

### SLE GWAS datasets and MHC imputation

For the association analysis of SLE, we used two independent genotype datasets from Korean populations, comprising 2,023 cases and 71,125 controls generating using Axiom Korean Chip^55^ (discovery GWAS dataset), and 348 cases and 4,203 controls generated using Illumina Omni BeadChip (replication GWAS dataset) after sample-level QC (**Supplementary Table 5**). In the discovery dataset, SNPs were filtered using a genotype call rate threshold of 0.05, a minor allele frequency cutoff of 0.005, and HWE threshold of 1×10^-6^ in cases and 1×10^-10^ in controls. Additionally, sample-level call rates, heterozygosity, population stratification, cryptic relatedness, and sex consistency were assessed for QC at the sample level. Detailed description of the replication dataset including the stringent QC criteria applied, have been provided elsewhere^5^. We extracted SNPs located in the extended MHC region (25-35 Mb on chromosome 6; hg38) from both datasets as for imputation inputs. The imputation was performed for each case-control dataset separately, using IMPUTE5^54^ and our MHC imputation reference panel.

### Calculation of correlation between HLA amino acid positions

As an approximate LD index, we estimated “lazy” r^2^ values to represent the correlation between two multi-residue amino acid positions within a single HLA gene. These estimates were derived by averaging the highest r^2^ values, where each maximum was the largest correlation between any residue at one position and all residues at the other position.

### Disease association analysis

The SLE association of each binary-encoded MHC variable (including each HLA classical allele, amino acid residue, *C4*-related haploid copy number, SNP, and indel, with an allele frequency > 0.5%) was assessed using logistic regression, incorporating imputed allelic dosage and covariates such as the top five PCs and sex. For the combined analysis using both datasets, a dataset-indicating dummy variable was additionally included as a covariate. To investigate the association of diploid copy numbers of *C4*-related elements, we estimated the diploid copy numbers (D) of *C4A*, *C4B* and HERV from the imputed dosages of haploid copy number variables using the formula 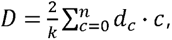 where 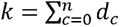. Here, *d_c_* represents the dosage of the “*T*” allele, indicating the presence of *c* haploid copies in an individual and is normalized by *k/2*, which is half the sum of the dosages of the “*T*” alleles for all possible haploid copies (ranging from 0 copy to *n* copies; *n*=3 for *C4A* and *C4B*, and *n*=4 for HERV). The total imputed copy number of *C4* is calculated as the sum of the diploid copy numbers of *C4A* and *C4B*.

To evaluate the multi-allelic effect of each amino acid position or *C4*-related elements on SLE risk, an LRT was conducted to compare a null logistic regression model including only covariates with a full model that additionally included multi-allelic variables. For each multi-residue amino acid position, the full model incorporated the dosages of amino acid residues and the same covariates, excluding the most common residue and any residues with a frequency below 0.5%. For *C4*, the full model included the diploid or haploid copy numbers of multiple *C4*-related elements (*C4A*, *C4B*, and/or HERV) along with the same covariates, and also included the dosages of G-group *HLA-DRB1* classical alleles, except the most common classical allele, as additional covariates to adjust for the multi-allelic effect of *HLA-DRB1*.

After the most significant variant in the MHC region was identified at a gene, a stepwise conditional regression analysis was performed, where each of biallelic and multiallelic variants in the MHC region was tested for the independent association with SLE risk, adjusting for the overall multi-allelic effects of the identified gene(s), as described previously^56^. This process was repeated until no additional variants reached the genome-wide significance threshold of 5×10^-8^. To account for the overall multi-allelic effects of HLA or *C4*, the dosages of G-group HLA classical alleles (excluding the most common allele and any alleles with a frequency below 0.5%) or the imputed diploid copy number of *C4A*, *C4B*, and HERV were added as covariates in the conditional regression models, respectively.

### Correlation analysis of plasma C4 protein level and *C4* copy numbers

In a subset of patients with SLE from the discovery GWAS dataset (n=1,284), plasma C4 concentrations were measured using immunoturbidimetric method with a normal reference range of 10-40 mg/dL, and disease activity was assessed with the SLE Disease Activity Index 2000 (SLEDAI). Linear regression analysis was performed to investigate the linear relationship between plasma C4 levels and copy numbers of *C4*-related elements, adjusting for disease activity, sex, and the top five PCs.

## Data Availability

All data except SLE GWAS datasets produced in the present study are available upon reasonable request to the authors. This individual-level genotype and phenotype data are protected and are not available due to data privacy laws.

## Data availability

Raw WGS data from reference batch #1 will be available as possible upon request and after approval from the Korean Genomics Center’s review board in UNIST. Raw WGS data from reference batch #2 will be available as possible upon request and after approval from KNIH. The SLE GWAS datasets were collected by Hanyang University. This individual-level genotype and phenotype data are protected and are not available due to data privacy laws.

## Code availability

The code of all tools used for analyses in this paper is publicly available and has been presented in the Methods section. Custom codes used to analysis are available at https://github.com/KimLabKHU/MHC-_reference_panel.

## Acknowledgements

We are grateful to all collaborators and study subjects for their participation in this study. This work was supported by the National Research Foundation (NRF) of Korea [NRF-2021R1A6A1A03038899, NRF-2022R1A2C2092164], the National Facility & Equipment Center [2023R1A6C101A009], the Hanyang University Institute for Rheumatology Research, the U-K BRAND Research Fund [1.200108.01] of UNIST, the Research Project Funded by Ulsan City Research Fund [1.200047.01] of UNIST, and the Ministry of SMEs and Startups (MSS, Korea) [P0016195, P0016193, P0016191; 1425156792, 1425157301, 1425157253; 2.220035.01, 2.220036.01, 2.220037.01]. Further funding was provided by the Korea Planning & Evaluation Institute of Industrial Technology with support from the Ministry of Trade, Industry and Energy in 2024 [RS-2024-00435468, Development and Dissemination of National Standard Technology].

## Authors’ contributions

SCB and KK designed the study. SJ and JB contributed to the collection and processing of WGS data for reference batch #1. DMS, SMK, YJK, and BJK contributed to the collection and processing of WGS data for reference batch #2. CYY and KK conducted all computational analyses with assistances from DMS, SMK, and SC. YTL and SJY conducted ddPCR experiments to quantify diploid copy numbers of *C4* elements. SYB, HSL, and SCB characterized patients with SLE and collected clinical data. SYB, HSL, YJK, BJK, SCB, and KK generated GWAS data. CYY, DMS, XY, YC, XZ, YJK, BJK, SCB, and KK interpreted the results. CYY and KK wrote the manuscript. All authors reviewed and approved the manuscript.

