## Supplementary Information for "Fine-mapping SLE-MHC associations revealed independent contributions of HLA missense variants and *C4* copy number variations"

**Supplementary Table 1.** ***C4*** **haploid copy number composition allocated to haplotype clusters**

| Cluster ID[a] | Haploid copy number | | | Notation[b] | Structural composition[c] | Frequency |
|---|---|---|---|---|---|---|
| | C4A | *C4B* | HERV | | | |
| 1 to 14 | 1 | 1 | 1 | A1+B1+L1 | AL+BS or AS+BL | 0.402 |
| 15 to 36 | 1 | 1 | 2 | A1+B1+L2 | AL-BL | 0.353 |
| 37 | 1 | 2 | 1 | A1+B2+L1 | AL+BS+BS or AS+BL+BS | 0.059 |
| 38 | 2 | 1 | 2 | A2+B1+L2 | AL+AL+BS or AL+AS+BL | 0.043 |
| 39 | 0 | 2 | 1 | B2+L1 | BL+BS | 0.038 |
| 40 | 3 | 1 | 4 | A3+B1+L4 | AL+AL+AL+BL | 0.032 |
| 41 | 2 | 0 | 2 | A2+L2 | AL+AL | 0.025 |
| 42 | 1 | 0 | 1 | A1+L1 | AL | 0.020 |
| 43 to 44 | 2 | 1 | 3 | A2+B1+L3 | AL+AL+BL | 0.019 |
| 45 | 0 | 3 | 3 | B3+L3 | BL+BL+BL | 0.007 |
| 46 | 1 | 1 | 0 | A1+B1 | AS+BS | 0.001 |
| 47 | 2 | 1 | 1 | A2+B1+L1 | AL+AS+BS or AS+AS+BL | 0.001 |

[a]The cluster #48 was deleted as no haplotypes were assigned to it based on cluster-specific likelihood estimates.
[b]The same notation used in Figure 3bAL, representing the initials and copy numbers of *C4A* (A), *C4B* (B), and HERV (L; long form).
[c]AL: HERV-inserted *C4A*; BL: HERV-inserted *C4B*; AS: HERV-free *C4A*; BS: HERV-free C4B.

**Supplementary Table 2.** **Hardy-Weinberg equilibrium analysis of *C4*-related CNVs**

***C4A***

| Diploid CN | 0 | 1 | 2 | 3 | 4 | 5 | 6 |
|---|---|---|---|---|---|---|---|
| Observed CN[a] | 4 | 183 | 1028 | 259 | 104 | 13 | 0 |
| Expected CN[b] | 7.40 | 172.26 | 1026.22 | 281.01 | 92.64 | 10.11 | 1.36 |
| *P*-value[c] | | | | 0.69 | | | |

***C4B***

| Diploid CN | 0 | 1 | 2 | 3 | 4 | 5 | 6 |
|---|---|---|---|---|---|---|---|
| Observed CN[a] | 21 | 268 | 1028 | 225 | 44 | 4 | 1 |
| Expected CN[b] | 17.85 | 267.95 | 1034.82 | 222.20 | 44.40 | 3.53 | 0.26 |
| *P*-value[c] | | | | 1.00 | | | |

**HERV**

| Diploid CN | 0 | 1 | 2 | 3 | 4 | 5 | 6 | 7 | 8 |
|---|---|---|---|---|---|---|---|---|---|
| Observed CN[a] | 0 | 3 | 474 | 671 | 312 | 93 | 33 | 5 | 0 |
| Expected CN[b] | 0.03 | 6.96 | 461.41 | 676.35 | 320.17 | 90.65 | 31.61 | 2.98 | 0.84 |
| *P*-value[c] | | | | | 0.90 | | | | |

[a]Observed CN denotes the true diploid copy number of *C4* elements, determined from whole-genome-sequencing data.
[b]Expected CN denotes the diploid copy number of *C4* elements, calculated from the frequencies of inferred haploid copy numbers under the assumption of Hardy-Weinburg equilibrium.
[c]The significance of the differences between observed and expected copy numbers was assessed using a simulated Fisher's exact test.
CN, copy number.

**Supplementary Table 3. Contingency table of diploid copy numbers of *C4* elements from leave-one-out cross-validation**

***C4A***

| Imputed CN[a] / True CN[b] | 0 | 1 | 2 | 3 | 4 | 5 | Sensitivity |
|---|---|---|---|---|---|---|---|
| **0** | 4 | 0 | 0 | 0 | 0 | 0 | 1.00 |
| **1** | 1 | 145 | 29 | 0 | 0 | 0 | 0.83 |
| **2** | 0 | 14 | 953 | 28 | 2 | 0 | 0.96 |
| **3** | 0 | 3 | 40 | 195 | 11 | 1 | 0.78 |
| **4** | 0 | 0 | 1 | 14 | 83 | 1 | 0.84 |
| **5** | 0 | 0 | 0 | 2 | 6 | 4 | 0.33 |
| **Precision** | 0.80 | 0.90 | 0.93 | 0.82 | 0.81 | 0.67 | |

***C4B***

| Imputed CN[a] / True CN[b] | 0 | 1 | 2 | 3 | 4 | 5 | 6 | Sensitivity |
|---|---|---|---|---|---|---|---|---|
| **0** | 11 | 8 | 0 | 0 | 0 | 0 | 0 | 0.58 |
| **1** | 2 | 205 | 48 | 0 | 0 | 0 | 0 | 0.80 |
| **2** | 0 | 23 | 948 | 22 | 2 | 0 | 0 | 0.95 |
| **3** | 0 | 2 | 31 | 183 | 3 | 0 | 0 | 0.84 |
| **4** | 0 | 0 | 2 | 1 | 41 | 0 | 0 | 0.93 |
| **5** | 0 | 0 | 0 | 0 | 2 | 2 | 0 | 0.50 |
| **6** | 0 | 0 | 0 | 0 | 0 | 0 | 1 | 1.00 |
| **Precision** | 0.85 | 0.86 | 0.92 | 0.89 | 0.85 | 1.00 | 1.00 | |

**HERV**

| Imputed CN[a] / True CN[b] | 1 | 2 | 3 | 4 | 5 | 6 | 7 | Sensitivity |
|---|---|---|---|---|---|---|---|---|
| **1** | 1 | 2 | 0 | 0 | 0 | 0 | 0 | 0.33 |
| **2** | 0 | 425 | 25 | 1 | 0 | 0 | 0 | 0.94 |
| **3** | 0 | 15 | 607 | 24 | 1 | 0 | 0 | 0.94 |
| **4** | 0 | 0 | 31 | 265 | 10 | 1 | 0 | 0.86 |
| **5** | 0 | 0 | 0 | 15 | 74 | 3 | 0 | 0.80 |
| **6** | 0 | 0 | 0 | 0 | 1 | 31 | 0 | 0.97 |
| **7** | 0 | 0 | 0 | 0 | 1 | 1 | 3 | 0.60 |
| **Precision** | 1.00 | 0.96 | 0.92 | 0.87 | 0.85 | 0.86 | 1.00 | |

[a]Imputed CN denotes the diploid copy number of *C4* elements calculated from the imputed dosages of haploid copy alleles using the reference panel that excluded the sample being imputed.
[b]True CN denotes the diploid copy number of *C4* elements determined from the whole-genome-sequencing data.
CN, copy number.

**Supplementary Table 4. Imputation accuracy of the reference panel for each haploid copy number via leave-one-out cross-validation**

| ***C4* element** | **Haploid copy number** | **Frequency** | **Sensitivity** |
|---|---|---|---|
| *C4A* | 0 | 0.068 | 0.86 |
| | 1 | 0.794 | 0.95 |
| | 2 | 0.109 | 0.77 |
| | 3 | 0.028 | 0.84 |
| *C4B* | 0 | 0.104 | 0.80 |
| | 1 | 0.795 | 0.94 |
| | 2 | 0.087 | 0.85 |
| | 3 | 0.013 | 0.90 |
| HERV | 0 | 0.002 | 0.67 |
| | 1 | 0.535 | 0.96 |
| | 2 | 0.398 | 0.92 |
| | 3 | 0.042 | 0.63 |
| | 4 | 0.023 | 0.94 |

**Supplementary Table 5. Characteristics of two independent GWAS datasets**

| GWAS dataset | Cases | Controls | Published | SLE criteria | GWAS array | Number of genotyped variants (QC-passed) | | $\lambda_{1000}$[b] |
|---|---|---|---|---|---|---|---|---|
| | | | | | | Total | MHC region (chr6:25-35Mb) | |
| Discovery dataset | 2,023 | 71,125 | expanded from our previous study (PMID:33272962) | 1997 ACR criteria | Axiom Korean Chip[a] | 479,193 | 4,974 | 1.025 |
| Replication dataset | 348 | 4,203 | PMID:25533202 | 1997 ACR criteria | Illumina Omni BeadChip | 520,210 | 5,615 | 1.047 |

[a]Details of the GWAS array are described in Moon *et al*. (PMID:30718733).
[b]Genomic inflation factor ($\lambda_{1000}$)
chr, chromosome.

**Supplementary Table 6. Significant associations between plasma C4 protein levels and the copy numbers of *C4* and HERV**

| Model | Variable | Effect (95% CI) | *P*-value |
|---|---|---|---|
| Plasma C4 level ~ *C4* CN + HERV CN + SLEDAI + Sex + PCs | *C4* CN | 6.50 (5.82 to 7.18) | 7.12E-69 |
| | HERV CN | -2.80 (-3.34 to -2.26) | 3.05E-23 |
| | SLEDAI | -0.48 (-0.60 to -0.36) | 2.87E-14 |
| | Sex | -1.30 (-2.96 to 0.35) | 1.22E-01 |
| Plasma C4 level ~ *C4A* CN + *C4B* CN + HERV CN + SLEDAI + Sex + PCs | *C4A* CN | 6.25 (5.41 to 7.10) | 2.64E-44 |
| | *C4B* CN | 6.69 (5.90 to 7.47) | 4.86E-57 |
| | HERV CN | -2.71 (-3.28 to -2.14) | 5.77E-20 |
| | SLEDAI | -0.48 (-0.60 to -0.36) | 3.01E-14 |
| | Sex | -1.33 (-2.98 to -0.32) | 1.16E-01 |

Linear regression was conducted on plasma C4 protein levels using imputed *C4*-related CNs and other variables, adjusting for the top five genotypic PCs.
CI, confidence interval; CN, diploid copy number; SLEDAI, systemic lupus erythematosus disease activity index; PCs, principal components.

**Supplementary Table 7. Highest correlation coefficient between *C4* haploid copy numbers and HLA alleles in the discovery GWAS dataset**

| | ***C4*** | | | ***C4A*** | | | ***C4B*** | | | **HERV** | | |
|---|---|---|---|---|---|---|---|---|---|---|---|---|
| | **Highest $r^2$** | **Which allele?** | **Allelic freq (%)** | **Highest $r^2$** | **Which allele?** | **Allelic freq (%)** | **Highest $r^2$** | **Which allele?** | **Allelic freq (%)** | **Highest $r^2$** | **Which allele?** | **Allelic freq (%)** |
| ***A*** | 0.018 | 24:02 | 12.3 | 0.044 | 24:02 | 12.3 | 0.048 | 33:03 | 5.7 | 0.033 | 24:02 | 12.3 |
| ***B*** | 0.331 | 07:02 | 2.0 | 0.366 | 07:02 | 2.0 | 0.112 | 15:07 | 0.5 | 0.339 | 07:02 | 2.0 |
| ***C*** | 0.080 | 07:02 | 4.8 | 0.095 | 07:02 | 4.8 | 0.106 | 07:01 | 1.3 | 0.131 | 07:02 | 4.8 |
| ***DPA1*** | 0.000 | 02:01 | 4.6 | 0.004 | 02:01 | 4.6 | 0.004 | 02:01 | 4.6 | 0.003 | 02:01 | 4.6 |
| ***DPB1*** | 0.043 | 04:02 | 5.1 | 0.053 | 04:02 | 5.1 | 0.069 | 09:01 | 1.2 | 0.036 | 04:02 | 5.1 |
| ***DQA1*** | 0.178 | 01:01 | 7.2 | 0.212 | 01:01 | 7.2 | 0.052 | 02:01 | 2.7 | 0.198 | 01:01 | 7.2 |
| ***DQB1*** | 0.217 | 05:01 | 5.7 | 0.248 | 05:01 | 5.7 | 0.114 | 06:09 | 1.2 | 0.240 | 05:01 | 5.7 |
| ***DRB1*** | 0.373 | 01:01 | 3.7 | 0.417 | 01:01 | 3.7 | 0.092 | 15:02 | 1.5 | 0.445 | 01:01 | 3.7 |

For each *C4* element and HLA gene, $r^2$ values were calculated from all possible pairs between HLA alleles (coded as 1 for presence and 0 for absence) and *C4*-related haploid copy numbers. The two-field HLA allele with the highest $r^2$ value among those of all pairs is listed along with its frequency.

**Supplementary Table 8. Interaction effects between *C4*-related CNVs and sex on SLE risk, adjusting for *HLA-DRB1* effects**

| SLE model | Term | Discovery GWAS dataset | | Replication GWAS dataset | | Combined[a] | |
|---|---|---|---|---|---|---|---|
| | | Effect (95% CI) | *P*-value | Effect (95% CI) | *P*-value | Effect (95% CI) | *P*-value |
| ~ *C4*+*C4*:Sex | *C4*:Sex | -0.02 (-0.23 to 0.19) | 0.085 | -0.01 (-0.64 to 0.63) | 0.984 | -0.02 (-0.22 to 0.18) | 0.865 |
| ~ *C4A*+*C4A*:Sex | *C4A*:Sex | 0.06 (-0.17 to 0.29) | 0.589 | 0.22 (-0.39 to 0.83) | 0.474 | 0.08 (-0.13 to 0.30) | 0.442 |
| ~ *C4B*+*C4B*:Sex | *C4B*:Sex | -0.11 (-0.35 to 0.14) | 0.401 | -0.29 (-1.05 to 0.47) | 0.453 | -0.12 (-0.36 to 0.11) | 0.310 |
| ~ HERV+HERV:Sex | HERV:Sex | -0.05 (-0.21 to 0.12) | 0.573 | 0.01 (-0.46 to 0.49) | 0.958 | -0.04 (-0.2 to 0.12) | 0.615 |

Logistic regressions were conducted on SLE using diploid-level *C4*-related CNVs risk with various interaction terms, adjusting for sex and genotype PC1-5, and conditioning on G-group *HLA-DRB1* alleles.

[a]In the combined analysis, a batch term (dataset #1 or dataset #2) was additionally included as a covariate in the model.

CI, confidence interval.

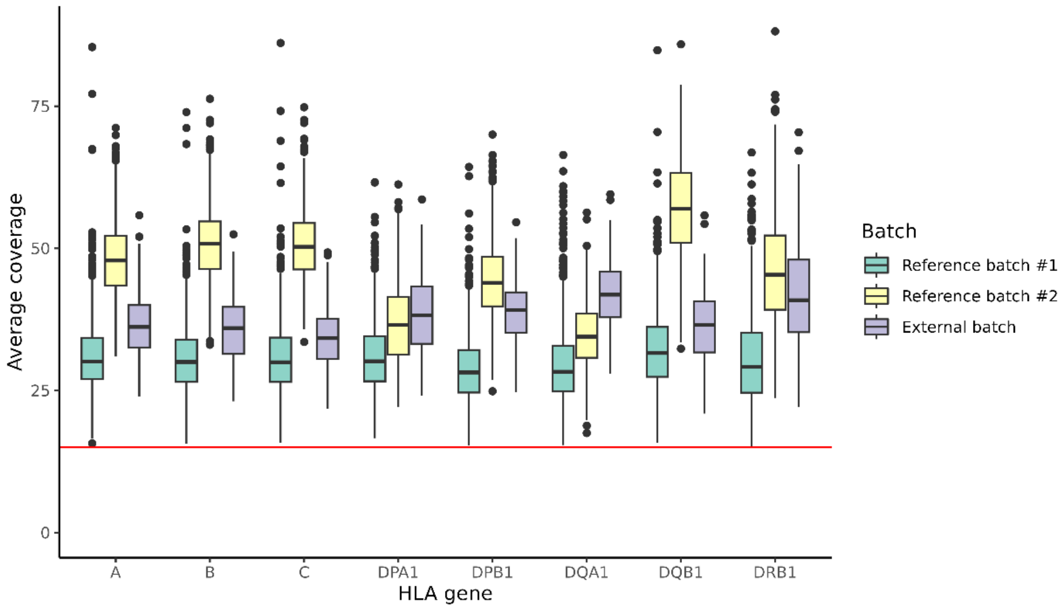


**Supplementary Fig. 1 | Sequencing coverage of HLA genes across two reference batches and one external batch.** Each box plot represents the average coverage for a specific HLA gene, with the red horizontal line indicating the quality control threshold at an average coverage of 15. Only samples exceeding this threshold for all HLA genes are shown and included in subsequent analyses.

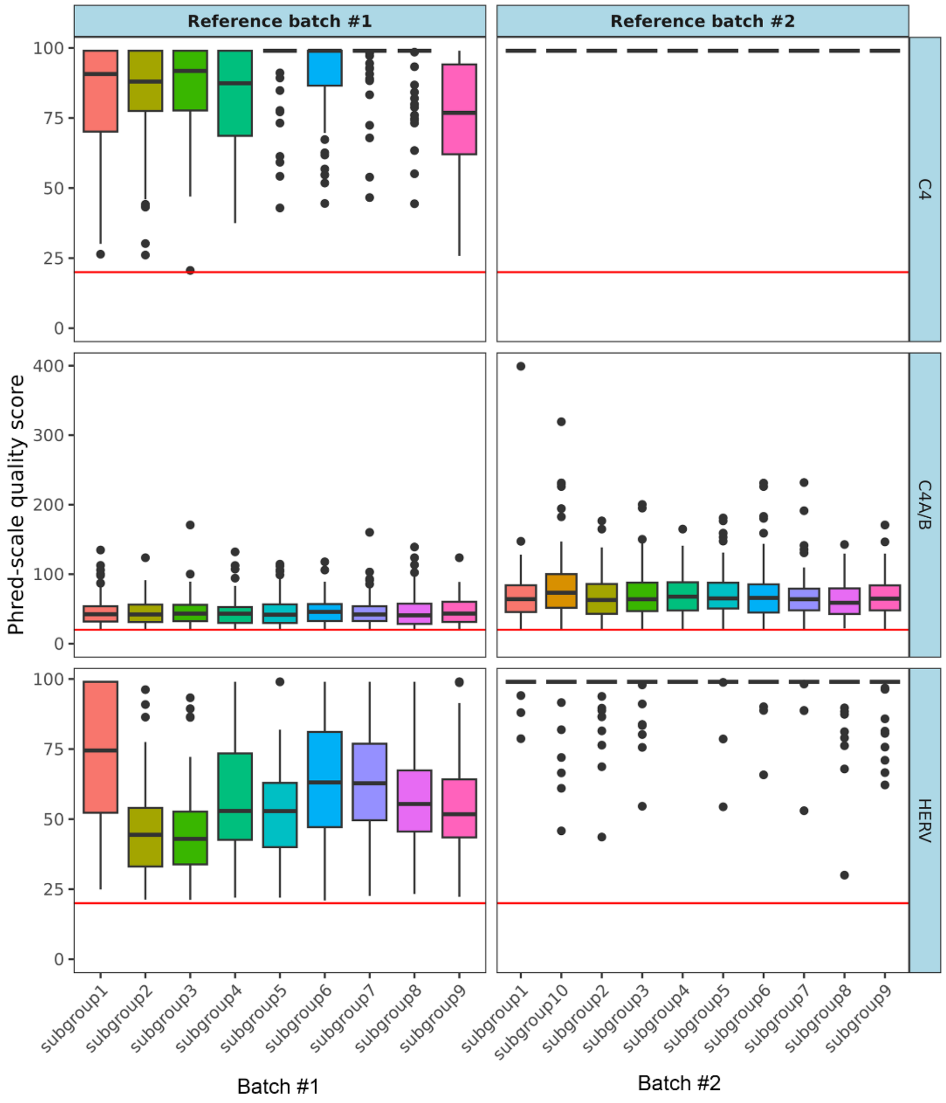


**Supplementary Fig. 2 | Phred-scale quality scores of diploid copy number calls in reference samples.** Boxplots display the quality scores for subgroups randomly divided within each reference batch due to limited computational capacity during program execution. A red horizontal line in each panel indicates the quality threshold set at 20 for sample quality control. Only samples that surpassed this threshold across all *C4* elements are plotted and included in subsequent analyses.

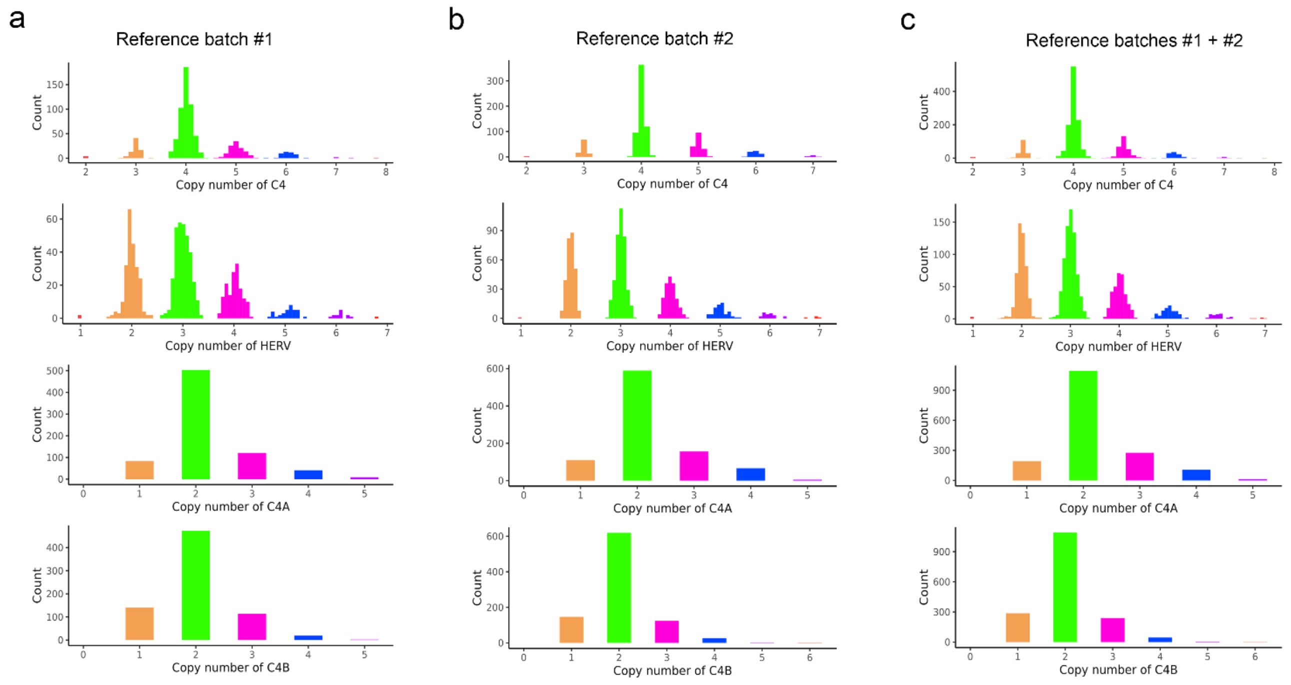


**Supplementary Fig. 3 |** **Distribution of diploid copy numbers across reference batches.** Diploid copy numbers of *C4* elements (*C4*, *C4A*, *C4B* and HERV) were determined using WGS data by Genome STRiP, as detailed in the Methods section. The distributions of copy numbers for these elements are plotted across **(a)** batch #1, **(b)** batch #2, and **(c)** the combined set. For *C4* and HERV, copy numbers were determined based on read depths mapped to their genomic regions. The estimated copy number values are clustered and color-coded by their corresponding copy numbers. For *C4A* and *C4B*, copy numbers were derived as integer values from the total *C4* copy number and the ratio of allele-specific reads mapped to *C4A* and *C4B*. Only samples with a Phred-scale quality score exceeding 20 for all *C4* elements were included in the distributions. No significant batch effects were observed in the copy number distributions of all *C4* elements between the reference batches.

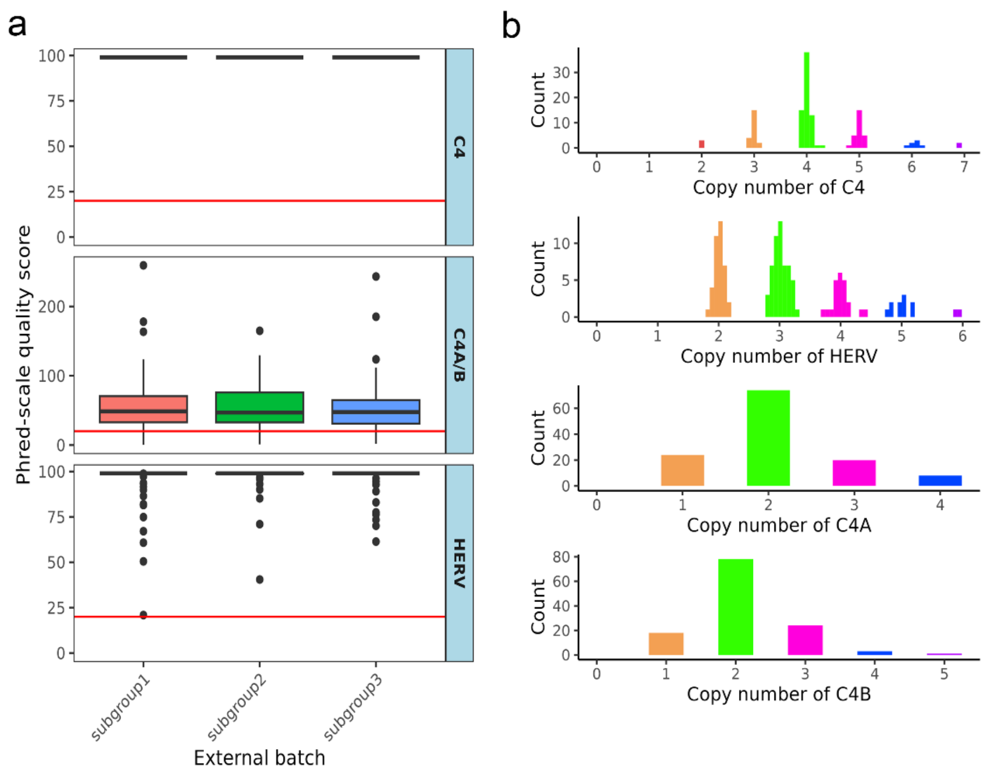


**Supplementary Fig. 4 | Distribution of diploid copy numbers of *C4* elements from WGS data of external validation samples. a.** Diploid copy numbers of *C4* elements (*C4*, *C4A*, *C4B*, and HERV) were determined using WGS data of 144 non-reference samples by Genome STRiP. Boxplots of Phred-scale quality scores for *C4*, *C4A/B* and HERV are displayed across subgroups that were randomly divided due to limited computational capacity during program execution. A red horizontal line in each panel indicates the quality threshold set at 20 for sample quality control. A total of 127 samples that surpassed this threshold across all *C4* elements are plotted and included in subsequent analyses. **b.** The distribution of identified copy numbers is presented for each *C4* element. For *C4* and HERV, copy numbers were determined based on read depths mapped to their genomic regions. The estimated copy values are clustered and color-coded by their corresponding copy numbers. For *C4A* and *C4B*, copy numbers were derived as integer values from the total *C4* copy number and the ratio of allele-specific reads mapped to *C4A* and *C4B*.

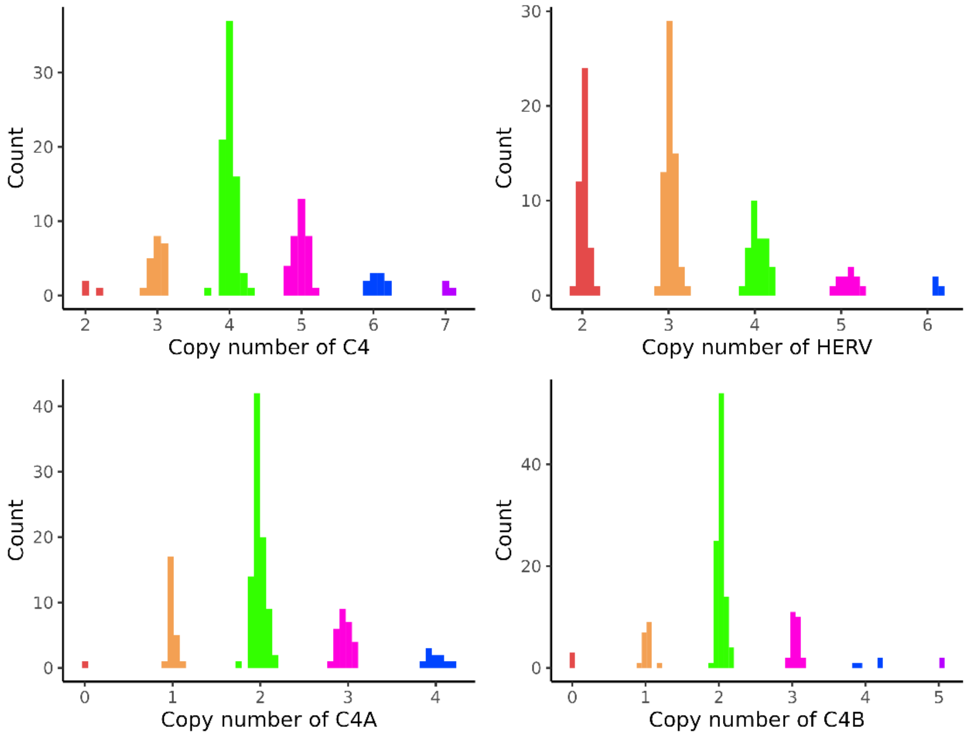


**Supplementary Fig. 5 | Distribution of diploid copy numbers of *C4* elements from ddPCR data of external validation samples.** Diploid copy numbers of *C4* elements (*C4*, *C4A*, *C4B*, and HERV) were determined using ddPCR data from 144 non-reference samples. Each sample was analyzed four times with four different probes to estimate the copy numbers for HERV-inserted *C4* (C4L), HERV-free *C4* (C4S), *C4A*, and *C4B*. Copy number estimates were obtained based on the ratio of target copies to reference copies (*RPP30*; two copies) and rounded to integer values. It was confirmed that in all samples, the sum of C4L and C4S copy numbers equaled the sum of C4A and C4B copy numbers. The distributions of copy estimates (unrounded) for all *C4* elements are color-coded by their corresponding integer-level copy numbers.

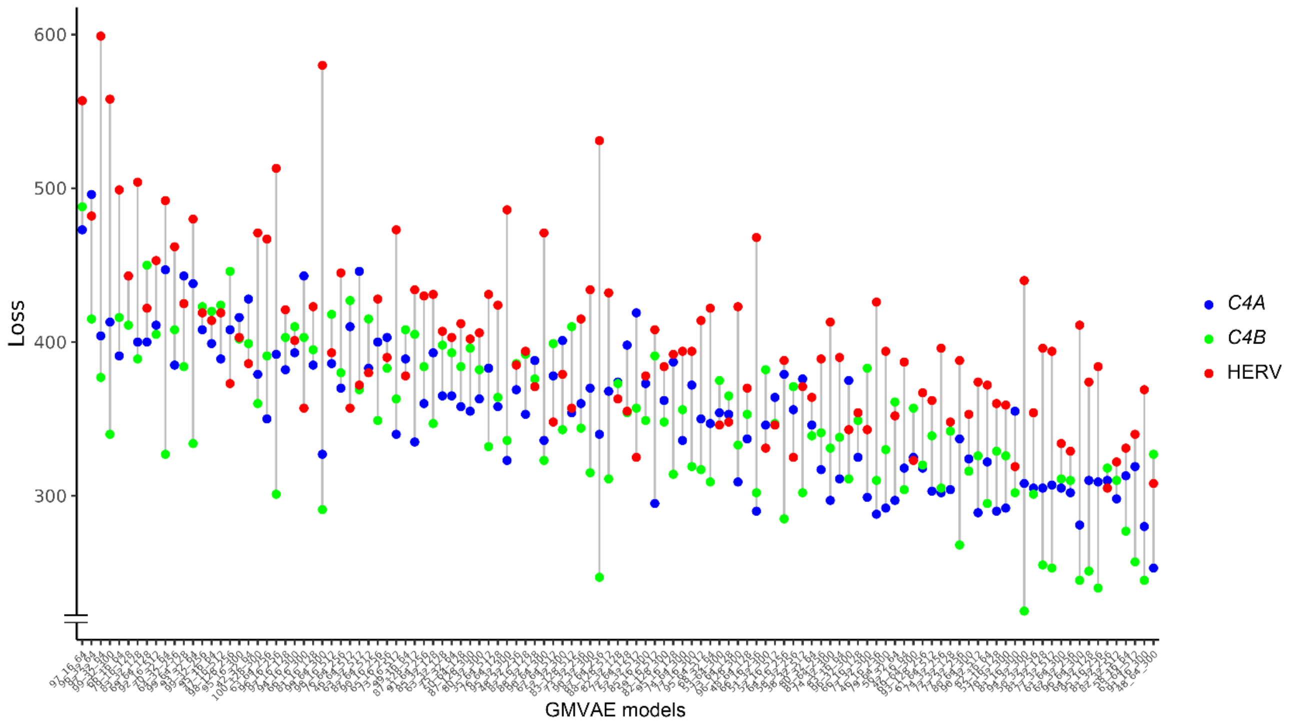


**Supplementary Fig. 6 | Evaluation of GMVAE models for unsupervised haplotype clustering.** Multiple GMVAE models within the HaploNet framework were evaluated to identify the optimal model for clustering haplotypes and subsequently segregating the diploid copy numbers of *C4A*, *C4B*, and HERV into haploid copy numbers. Models were defined by varying combinations of the numbers of hidden layer nodes (64, 128, 256, 300, 512), latent variables (16, 32, 64, 128), and clusters (1 to 100). For each GMVAE model, we allocated the most likely copy numbers of *C4* elements to each haplotype cluster using a cyclic coordinate descent method (see the Methods section). Only models that successfully allocated all haploid copy numbers for *C4A* (= 0,1,2,3), *C4B* (= 0,1,2,3), and HERV (= 0,1,2,3,4) into clusters were included for performance assessment. Model performance was measured by the average loss, representing the discrepancy between model cluster-based inferred diploid copy numbers and true diploid copy numbers for *C4A* (blue), *C4B* (green), and HERV (red). The model parameter combinations are represented on the x-axis as "(cluster number)_(latent variable number)_(hidden layer node number)". The model with 48 clusters, 64 latent variables, and 300 hidden layer nodes was selected as the best-performing model, achieving the lowest average loss.

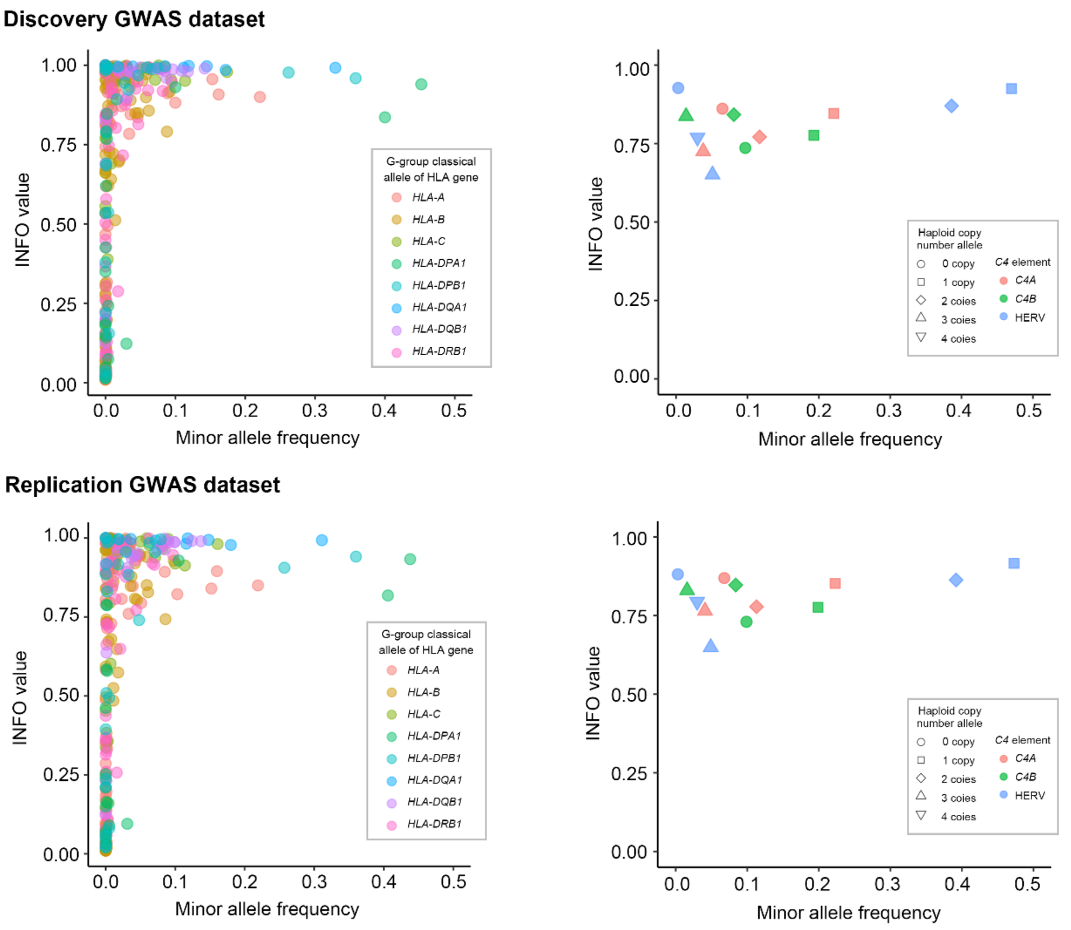


**Supplementary Fig. 7 | Imputation quality scores for *C4* and HLA variants in SLE GWAS datasets.** Imputation quality scores (INFO; from IMPUTE5) are plotted for G-group HLA classical alleles (left) and *C4*-related haploid copy number alleles (right) according to allele frequencies across the GWAS datasets.

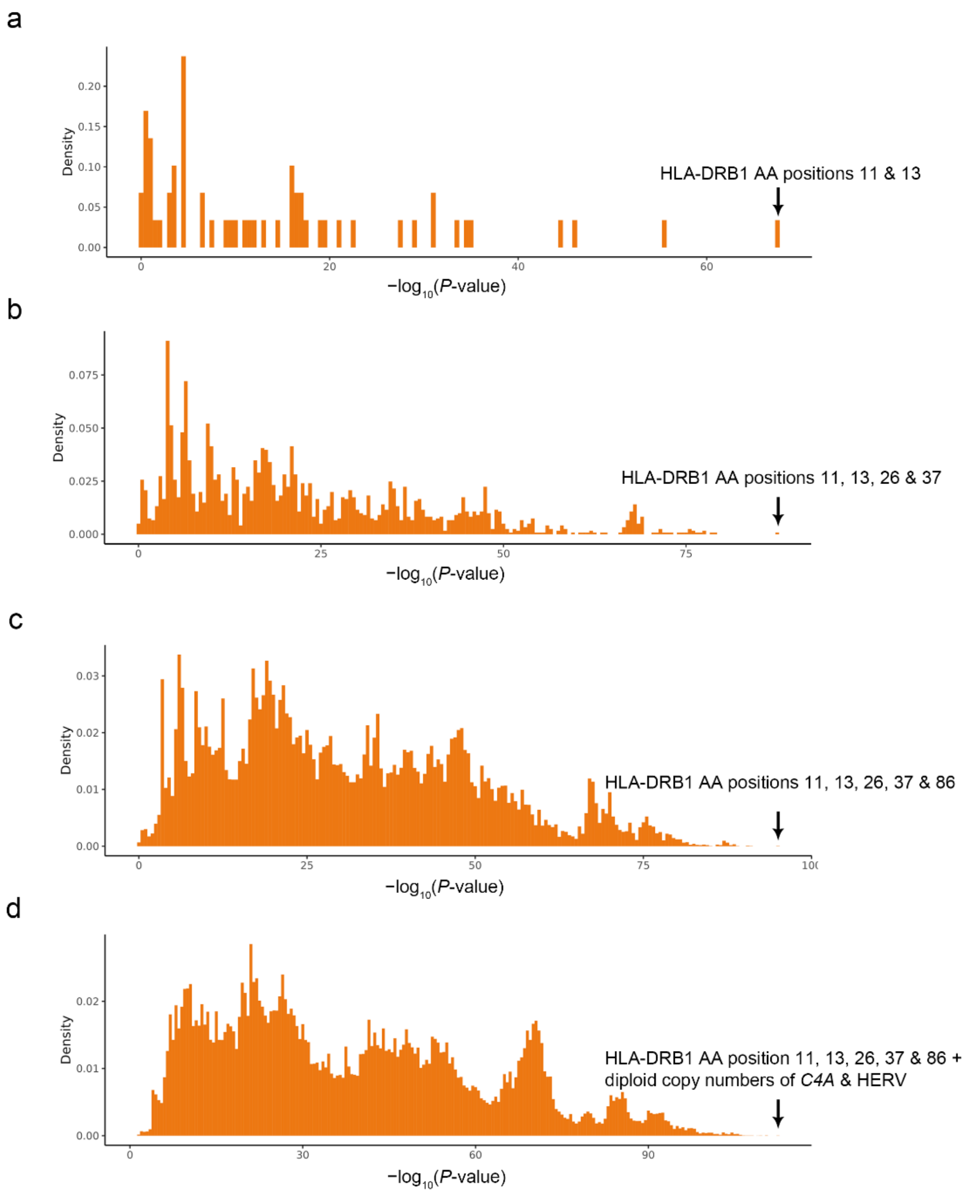


**Supplementary Fig. 8 | $P_{LRT}$ values for SLE associations explained by key variants within empirical distributions using SLE GWAS datasets. a.** The distribution of $P_{LRT}$ values from SLE association models, each incorporating a single association position from HLA-DRB1, identified the best model, which included a single association signal from positions 11 and 13, treated as a single unit due to their strong correlation. **b.** For models including two association signals derived from all potential combinations of HLA-DRB1 amino acid positions, the optimal model included positions 11, 13, 26, and 37, with positions 26 and 37 considered to share a single association signal due to their relatively high correlation and comparable contributions to SLE risk (see Results section). **c.** When three association signals from HLA-DRB1 amino acid positions were considered, position 86 emerged as an additional key contributor to the best-fitting model. **d.** Models that combined all possible combinations of three random *HLA-DRB1* positions with two *C4* elements (among *C4A*, *C4B*, HERV copy numbers) confirmed that the variants identified through our fine-mapping analysis provided the most robust explanation for SLE risk.
AA, amino acid.

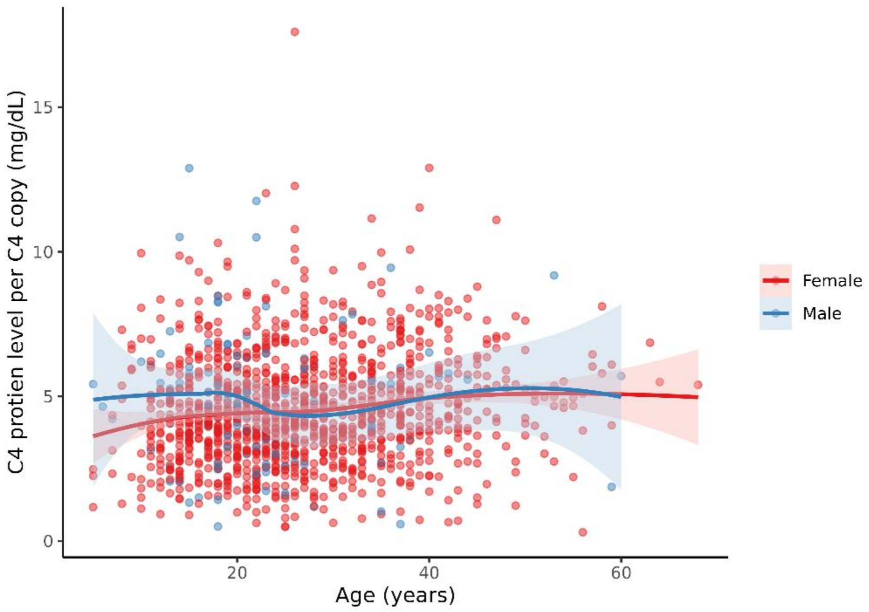


**Supplementary Fig. 9 | Plasma C4 protein levels per *C4* copy in SLE patients.** Plasma C4 protein levels, normalized by individual *C4* copy numbers, are plotted against age for 103 men (blue) and 1,181 women (red) from the SLE patient cohort. Locally estimated scatterplot smoothing (LOESS) is used to display the trend, with shaded areas representing 95% confidence intervals from the LOESS fit.
